# Opportunistic Assessment of Ischemic Heart Disease Risk Using Abdominopelvic Computed Tomography and Medical Record Data: a Multimodal Explainable Artificial Intelligence Approach

**DOI:** 10.1101/2021.01.23.21250197

**Authors:** Juan M Zambrano Chaves, Akshay S Chaudhari, Andrew L Wentland, Arjun D Desai, Imon Banerjee, Robert D Boutin, David J Maron, Fatima Rodriguez, Alexander T Sandhu, R Brooke Jeffrey, Daniel Rubin, Bhavik Patel

## Abstract

Current risk scores for predicting ischemic heart disease (IHD) risk—the leading cause of global mortality—have limited efficacy. While body composition (BC) imaging biomarkers derived from abdominopelvic computed tomography (CT) correlate with IHD risk, they are impractical to measure manually. Here, in a retrospective cohort of 8,197 contrast-enhanced abdominopelvic CT examinations undergoing up to 5 years of follow-up, we developed improved multimodal opportunistic risk assessment models for IHD by automatically extracting BC features from abdominal CT images and integrating these with features from each patient’s electronic medical record (EMR). Our predictive methods match and, in some cases, outperform clinical risk scores currently used in IHD risk assessment. We provide clinical interpretability of our model using a new method of determining tissue-level contributions from CT along with weightings of EMR features contributing to IHD risk. We conclude that such a multimodal approach, which automatically integrates BC biomarkers and EMR data can enhance IHD risk assessment and aid primary prevention efforts for IHD.

## Introduction

Ischemic heart disease (IHD) is the leading cause of global mortality and among the top causes of morbidity. In 2019, it was responsible for over 9 million deaths worldwide and the loss of more than 180 million disability-adjusted life years (http://ghdx.healthdata.org/gbd-results-tool). Preventive treatments including lifestyle modifications and pharmacologic interventions (e.g., cholesterol lowering and antiplatelet medications) can be guided by risk assessment. The Framingham coronary heart disease risk score (FRS) and the Pooled Cohort Equations (PCE) are commonly utilized risk estimation methods for IHD and atherosclerotic cardiovascular disease, respectively^1,2^. The FRS uses demographic risk factors and cholesterol values to predict 10-year IHD risk in individuals aged 30-74 years old without known IHD at baseline examination. The PCE were developed to model 10-year risk of major atherosclerotic cardiovascular disease events, including fatal and nonfatal IHD as well as fatal and nonfatal stroke. These risk scores have been used as a standard for IHD risk assessment in current clinical practice guidelines and policy recommendations, including the most recent American College of Cardiology/American Heart Association guideline on primary prevention of cardiovascular disease^3^.

Validation of both risk scores has shown varying performance depending on the subpopulation analyzed. Performance is typically reported as a c-statistic value, which corresponds to the proportion of case-control pairs in which a higher risk is assigned to the case (a measure of discrimination). Previously reported c-statistic values for the FRS and PCE are modest with typical ranges of .66-.76 and .68-.76, respectively,^4^ showing room for improvement. Thus, the discovery of additional biomarkers that improve or independently inform the predictive power of these existing models has been the objective of multiple recent research endeavors^5,6^.

Imaging biomarkers derived from computed tomography (CT) have shown promise in the assessment of cardiovascular risk. For example, the coronary artery calcium (CAC) score measures the extent of plaque in the coronary arteries from coronary CTs, and is an important tool for IHD risk stratification^7,8^. Although CAC scoring is a strong independent predictor of cardiovascular events^9^, the integration of both clinical factors (e.g. FRS) and imaging factors (e.g. CAC score) has been shown to significantly improve prediction of major cardiac events and all-cause mortality (compared to clinical or imaging metrics alone)^10,11^. Other studies have combined metrics from coronary CT angiography with blood biomarkers such as high-sensitivity cardiac troponin to successfully improve upon current measures of cardiovascular risk^12,13^. All of these specialized methods apply to a subset of patients already being assessed for cardiovascular risk.

Alternatively, abdominopelvic CTs contain body composition (BC) imaging biomarkers for atherosclerotic cardiovascular disease, such as hepatic steatosis^14^, low muscle mass^15^, an increased ratio of visceral to subcutaneous adipose tissue (VAT/SAT)^16^, and abdominal aortic calcification^17^. Notably abdominopelvic CTs are acquired almost twice as often than the CT scans that image the heart or coronary vessels, such as non-contrasted chest CT and coronary CT^18,19^. According to the National Hospital Ambulatory Care Survey (https://bit.ly/2SL6957), in 2016 over 10 million abdominopelvic CTs were acquired in the US during emergency department visits alone, often in relation to abdominal pain—the most common principal reason for visiting an emergency department^20^. Within abdominopelvic CTs, these biomarkers could be measured during such routine imaging procedures without resulting in additional costs or radiation exposure, referred to as opportunistic imaging^21^. However, the current clinical workflow and volume of imaging is not well-suited to allow practical utilization of the additional resources required to manually extract measurements of imaging biomarkers^22^. Consequently, despite the potential value, cardiovascular risk is not routinely assessed upon abdominopelvic CT acquisition, thereby missing opportunities for early disease detection and prevention.

In this work, we developed IHD risk assessment models that use automatically measured imaging features from abdominopelvic CT examinations in combination with information from the patient’s EMR. We evaluated the benefit of extracting BC imaging biomarkers from an axial slice at the level of the third lumbar vertebra (L3) in addition to traditional PCE metrics. We also developed an IHD risk assessment tool using the raw L3 slice image in an end-to-end manner using deep learning. We further developed methods to quantitatively assess the contribution of imaging features to the model prediction, aggregated at the tissue level. Finally, we combined features derived from the EMR in addition to the L3 slice, yielding the greatest risk prediction performance, and interpret the individual contribution of clinical features.

## Methods

### Study population

Following Stanford University institutional review board approval, we identified an initial cohort of 37,708 contrast-enhanced abdominopelvic CTs performed between January 2013 and May 2018 on individuals who presented to our tertiary center emergency department for abdominal pain. We included images with 1.0 or 1.25 mm axial spacing, from individuals 18 years of age or older with at least one documented clinical encounter in the year prior to and at least 1-year immediately following the acquisition of the image. For each patient, data from previous medical encounters were obtained. All demographic information (birthdate, gender, race/ethnicity), along with vital signs, body mass index, International Classification of Disease, 10^th^ edition codes (ICD10), Current Procedural Terminology (CPT) codes, laboratory results and prescriptions were extracted. We labeled individuals who had an ICD10 diagnosis code for Ischemic Heart Diseases (I20-I25) in the follow-up period after the image acquisition as IHD positive and those that did not have the code as negative. ICD codes have been found to have high sensitivity and specificity in identifying IHD in prior studies^23,24^. Since our goal was to identify new IHD patients that may not otherwise be detected, we excluded images from individuals with any diagnosis of IHD prior to and at the time of the image acquisition. We defined two cut-off periods for follow-up, 1 year and 5 years, establishing two cohorts representing individuals who either develop IHD or have follow-up within those time frames.

From each CT volume, we automatically identified the slice at L3 using a previously published convolutional neural network (CNN) algorithm^25^, manually verifying correctness for each case. The L3 slice was chosen as it is the most common reference location for BC analysis^26–29^. The process to select the final cohorts of patients is shown in Supplemental Figure 1. We excluded images with artifacts that obscured the L3 level (e.g., spinal instrumentation), those that had anatomical variations in the image (e.g., scoliosis) that precluded the assignment of a single slice to the L3 level, those that did not contain the L3 slice in the field of view and those obtained within the same 6 month window as an already included image. For each cohort, we used random sampling stratifying on outcome labels to divide patients in the dataset into training and test datasets representing 80% and 20% of the images respectively for IHD risk estimation and model creation.

### Segmentation Model

A total of 400 axial L3 slices obtained exclusively from the training set and manually labeled were used during model tuning and evaluation. Manual segmentation of muscle, SAT, VAT, and bone were performed semi-automatically with CoreSlicer^30^, a free online tool, using attenuation thresholds and manual adjustments as needed (AW, 5 years of experience). 320 images were randomly selected for segmentation training, 40 for validation, and 40 for testing. Segmentation performance on the 40 test sets was determined using segmentation accuracy metrics, namely the Dice coefficient and root-mean-squared coefficient-of-variation. The Dice coefficient is calculated as two times the ratio of the intersection between the ground-truth and segmented image masks to the sum of the number of pixels in each mask. A Dice score of 1 indicates a perfect segmentation. The variations between the manual and automated segmentation approaches on the tissue-wise HU and cross-sectional area were also evaluated.

Given that BC metrics from manual segmentations have been correlated with cardiovascular risk, we trained a 2.5D U-Net CNN to perform BC analysis for segmenting regions of muscle along with VAT and SAT within an abdominopelvic CT slice^31^. The inputs to the 2.5D network were individual 2D axial CT slices at the L3 level with three different window and level (W/L) settings for maximizing tissue contrast. The W/L settings that were used included a soft tissue window (W/L = 400/50 Hounsfield units [HU]), bone window (W/L = 1800/400 HU), and a custom window (W/L = 500/50 HU). After applying the appropriate windowing, each of the channels was normalized to values between 0 and 1.

The U-Net utilized 6 convolution levels (each with two convolutional operators, both followed by a rectified linear unit activation, followed by batch normalization) for the encoder and decoder^32^. The number of U-Net features per layer increased quadratically from 32 to 1024. The dimensions of the convolutional kernels were 3×3, while that of the maximum pooling operator was 2×2. A softmax activation was used as the final layer in the CNN along with a weighted soft Dice loss function to account for class-imbalance amongst the segmented tissues. The U-Net hyperparameters had previously been optimized for medical imaging segmentation^32^. A weighting factor of 8 was used for muscle during loss computation. All network weights were randomly initialized using the He et al initialization scheme^33^.

Training was performed with the Adam optimizer with default parameters ^β^1 0.9 ^β^2 of .999, with a learning rate schedule that included a base learning rate of 1e-3 and the learning rate being reduced by 0.8 for every epoch to a minimum value of 1e-8. The network was trained nominally trained for 130 epochs with an early stopping criterion of a minimum change in loss of 1e-5 and a patience of 8 epochs. The batch sizes for training, validation, and testing were chosen to be 10, 33, and 80 respectively for maximizing GPU memory. Training was performed using a Tensorflow 1.14 on an NVIDIA Titan Xp GPU.

We used the segmentations generated by the U-Net model and determined average muscle radiodensity in HU and the VAT/SAT cross-sectional area ratio. We trained a model (L2 logistic regression) using ten-fold cross validation on the training sets to predict IHD outcome at 1 and 5 years using these two features. We refer to this as the segmentation only model.

### Imaging Only Model

We trained a CNN as a feature extractor to predict the risk of IHD using a single axial slice at the L3 level, using an EfficientNet-B6 architecture^34^. EfficientNets were designed to balance the scaling of network width, depth and image size, thus producing state-of-the-art results in conventional image classification tasks with smaller and faster models as compared to traditionally used feature extractors, such as ResNet50^35^. The 512×512 pixel grayscale L3 slice was clipped to contain values from −1000 to 1000 HU, represented as an unsigned integer, replicated thrice to produce a 3×512×512 image, and resized to 3×528×528 to be input into the network. The initial EfficientNet-B6 model weights were obtained from a pre-trained model optimized for ImageNet classification performance (https://pypi.org/project/efficientnet-pytorch/)^36^. The final fully-connected layer was replaced with one corresponding to a binary outcome, and the model weights were fine tuned. The tuning of the weights was performed with a cross-entropy objective using a random selection of 80% of the training set, reserving 20% for validation. Training was performed for multiple epochs until no improvement in validation loss or Area Under the Receiver Operating Characteristic (AUROC) was observed. A batch size of 8 and an Adam optimizer^37^ was used with default parameters ^β^1 0.9 ^β^2 of .999 and a constant learning rate of 1e-5 and 1e-6 for the 1 and 5-year cohorts, respectively. Model training was carried out using Pytorch 1.1 on an NVIDIA Titan Xp GPU.

We compared training only the final layer as opposed to training all of the model weights, using additional image augmentations such as rotations of up to 3° and pixel shifting of up to 5 pixels during training, and using a focal loss function assigning higher weights to IHD cases. We chose the final network architecture and training strategy described above as it achieved the highest AUROC in the validation stage (Supplemental Table 1).

### Clinical Only Model

We used the data extracted from the EMR to produce features to develop a predictive model. Demographic data used were age at time of scan and gender. In this initial approach, we did not include race/ethnicity as a feature because of its limited accuracy in medical records^38^, in addition to obtaining no benefit in discrimination performance in preliminary results when including it as a covariate. From the EMR within one year prior to image acquisition, we obtained vital signs (blood pressure, heart rate, respiratory rate, oxygen saturation, temperature), body mass index, and relevant laboratory results (total, low-density lipoprotein, high-density lipoprotein cholesterol, triglycerides, fasting glucose and hemoglobin A1c). In multiple clinical encounters, vital signs and laboratory results were combined using an exponential weighting average, with each weight inversely proportional to the difference in time between the data point acquisition and the image acquisition. The number of times a vital sign or lab result was reported was also used as a feature. With the exception of select PCE covariates (low-density lipoprotein, high-density lipoprotein cholesterol), no imputation strategy was used for missing values.

To avoid sparsity, we grouped ICD10, CPT and medications based on their underlying ontology. Namely, we grouped ICD10 codes (https://bioportal.bioontology.org/ontologies/ICD10) by blocks and CPT Category I codes (https://bioportal.bioontology.org/ontologies/CPT) at the H2 level. Irrespective of dose and frequency, the active substance of prescribed medications was mapped to RxNorm and subsequently to the second level of the Anatomical Therapeutic Code (https://bioportal.bioontology.org/ontologies/ATC), corresponding to the therapeutic subgroup. In all, each patient image was represented using a 434-dimensional vector. The final clinical features used and their descriptions are listed in Supplemental Table 2.

The predictive model used for predicting IHD risk from EMR features was designed using XGBoost, an optimized gradient-boosting machine learning system^39^. In gradient boosting, an ensemble of weak learners is iteratively constructed by greedily adding estimators that fit the previous residual. In doing so, gradient boosting algorithms can perform successfully across a wide variety of predictive tasks, often outperforming traditional models such as logistic regression or support vector machines. We chose XGBoost for its robust performance in predictive modeling, and for its capacity to handle missing data, which other gradient boosted methods like AdaBoost lack. Optimal parameters for training the model were established using ten-fold cross-validation on the training set.

### Fusion models

To further identify the potential benefits of using imaging BC features as predictors of IHD risk, we constructed three models to fuse imaging and clinical data. In the first fusion, we concatenated the features used by the PCE with the average muscle radiodensity and the VAT/SAT ratio (*PCE + Segmentation model*), the latter two measurements obtained by using our automated segmentation model. In the second fusion, we combined the risk output from our EfficientNet-B6 model with the risk output from our medical record model using stacking with L2 logistic regression (*Imaging + Clinical model*). In the third fusion, we combined the risk output from the imaging only, clinical only and segmentation only models (Imaging + Clinical + Segmentation model). In all fusion cases, we performed a hyperparameter search in 10-fold cross-validation in the training set. The clinical model and fusion models were trained using scikit-learn 0.23 (https://scikit-learn.org/) in Python 3.6.

### Interpretation of model performance

Two baseline models currently employed in clinical practice that estimate 10-year cardiovascular risk were used as a reference, namely the FRS^1^ and the PCE^2^. We studied the performance of the FRS as it directly models risk of hard IHD events. Despite the PCE including other atherosclerotic cardiovascular disease outcomes, we also included them as a baseline given their current use in clinical practice guidelines. Since several subjects were missing covariates necessary for FRS and PCE calculation (Supplemental Table 3), these values were imputed using median imputation to allow for a baseline risk calculation for all individuals in the study. In addition, we examined the performance of all models in the subpopulations with available/missing PCE covariates (Supplemental Table 5).

### Attribution analysis

With the aim of allowing for interpretability of the fusion model, we examined the contributions of individual features in both the imaging and clinical models.

For the imaging model, we developed a new tissue saliency interpretation tool as described previously to evaluate the tissues that had a large contribution to the final prediction outcome. We first calculated the derivative, *w*, of the IHD class score at the final layer of our EfficientNet-B6 model, *S*_IHD_, with respect to each input pixel, *I*_*ij*_ in the image *I*^40^. That is,0

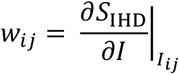

Using our segmentation model, each pixel was assigned a specific tissue class, *t*. In our particular case, this corresponded to either muscle, VAT, SAT, other body tissues or background. We obtain the observed normalized tissue saliency, *S*^*O*^, for a particular tissue, as:

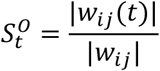

That is, the L1 norm of saliency of a particular tissue divided by the L1 norm of the total saliency. We contrast this observed saliency value with the expected tissue saliency,*S*^E^, which we define as:

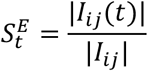

i.e. the proportion of pixels in the image belonging to the tissue *t*. We compared the proportion of observed vs. expected tissue saliency across the dataset by averaging the values for each image, 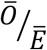

To assign specific tissue labels for each pixel, we first obtained a binary mask identifying the patient’s body by removing the background and CT bed using traditional image processing methods^41^. Specific tissue labels for fat and muscle within the body were automatically assigned using our segmentation CNN. Tissues within the body mask not belonging to VAT, SAT, or muscle were assigned to the *Other Tissues* class. All other pixels were considered the background.

To examine the relative contribution of each clinical feature to the prediction decision, we determined the SHapley Additive exPlanations (SHAP) values for each feature for each individual. SHAP values are an additive metric of feature importance that quantify the change in expected model prediction conditioned on a feature value^42^. In other words, they are a measure of how much the model prediction changes given a value for a particular feature. We used SHAP values to interpret the relative contribution of our clinical features to the final model classification output.

### Statistical analysis

The AUROC was used as the primary metric to compare and select models during training. The AUROC is equivalent to the c-statistic in the case of binary classification, as is traditionally reported in the cardiovascular risk assessment literature. In addition, we measured the Area Under the Precision Recall Curve (AUCPR), which is more informative in the case of imbalanced datasets, such as this one^43^. For both metrics, 95% confidence intervals were obtained using the stratified bootstrap method. We also report the sensitivity, specificity, positive and negative predictive values of our models at a sample threshold defined using Youden’s index.

Statistical analyses were carried out using SciPy 1.3^44^. Comparisons between AUROC values were carried out using the DeLong method^45^. Comparisons between AUCPR values were established using the stratified bootstrap method. Observed and expected tissue saliency values were compared using paired t-tests. All tests performed were two-tailed. An ^α^value of 0.05 was used to determine statistical significance.

## Results

### Final Patient Cohort

We collected a dataset of 8,197 CT images of individuals with at least 1-year of follow-up, with a subset of 1,762 images of 1,686 individuals with at least 5-years of follow-up. The average (interquartile range) length of follow-up was 3.6 (2.2) years. For each individual, data available in the EMR in the year before the scan acquisition was obtained. With 1 and 5 year follow-up after CT scan acquisition, a new IHD diagnosis was identified in 358 (4.5%) and 446 (25.4%) of individuals. Because sampling was performed in a stratified fashion, the prevalence in the training and test sets is equal for both cohorts. The average (standard deviation) age at time of scan in the dataset was 51.7 (17.5) years, with 40.5% of CT exams in men. Additional demographic characteristics of both cohorts along with PCE covariates and BC metrics are in Supplemental Table 3.

The L3 slice was correctly labeled in 8,156 (99.5%) of cases. In the 41 incorrect cases, the predicted L3 slice was 137 ^±^ 126 mm away from the correct location (mean ^±^ standard deviation). Incorrect localization typically occurred on CT exams with additional anatomical coverage, such as scans also including the chest or lower extremities. Otherwise, the automatically selected slice was at the L2 or L4 level, immediately adjacent to the L3 level.

The performance of the models on the held-out test set is reported as follows.

### Traditional IHD Risk Assessment Model Performance

The PCE outperformed the FRS in 1-year IHD prediction AUROC (.75 vs .71; *P*=.04), performing similarly in terms of AUCPR (.12 vs .09; *P*=0.06), and performed comparably in the case of 5-year IHD prediction, with AUROC .73 vs .71 (*P=*.24) and AUCPR .41 vs .40 (*P=*.73) respectively (Table 1). The ROC and precision-recall curves for the PCE are shown in Figure 2 and compared with the FRS and other proposed models in Supplemental Figure 2. Sensitivity, specificity, positive and negative predictive values for PCE at two clinically relevant cut-offs are shown in Supplemental Table 4.

**Table 1.** Proposed model performance in comparison to Pooled Cohort Equations (PCE) and Framingham Risk Score (FRS) as measured by area under receiver operating characteristic (AUROC) and precision-recall (AUCPR) curves. 95% confidence intervals (CI) and P values were obtained using the DeLong method for AUROC and the bootstrap method for AUCPR. Reported *P* values correspond to comparisons with the PCE.

| Model | 1y AUROC (95% CI) | P | 1y AUCPR (95% CI) | P | 5y AUROC (95% CI) | P | 5y AUCPR (95% CI) | P |
| --- | --- | --- | --- | --- | --- | --- | --- | --- |
| FRS | .71 (.67-.76) | **.04** | .09 (.07-.12) | .06 | .71 (.66-.76) | .24 | .40 (.35-.48) | .73 |
| PCE | .75 (.71-.81) | - | .12 (.10-.17) | - | .73 (.69-.78) | - | .41 (.36-.48) | - |
| Segmentation | .70 (.65-.74) | .10 | .08 (.07-.10) | .08 | .73 (.68-.78) | .85 | .43 (.38-.51) | .64 |
| PCE+Segmentation | .76 (.71-.81) | .68 | .12 (.10-.15) | .90 | .74 (.70-.79) | .45 | .43 (.38-.51) | .41 |
| Clinical only | .76 (.72-.81) | .57 | .12 (.10-.17) | .98 | .84 (.80-.87) | **<.001** | .64 (.58-.72) | **<.001** |
| Imaging only | .74 (.70-.78) | .70 | .10 (.08-.15) | .55 | .81 (.76-.85) | **.02** | .64 (.57-.71) | **<.001** |
| Imaging+Clinical Fusion | **.77 (.73-.81)** | .38 | **.13 (.10-.19)** | .73 | **.86 (.82-.90)** | **<.001** | **.70 (.63-.77)** | **<.001** |
| Imaging+Clinical+Segmentation Fusion | .74 (.70-.79) | .74 | **.13 (.10-.18)** | .78 | **.86 (.82-.89)** | **<.001** | **.70 (.63-.77)** | **<.001** |

**Figure 1.**
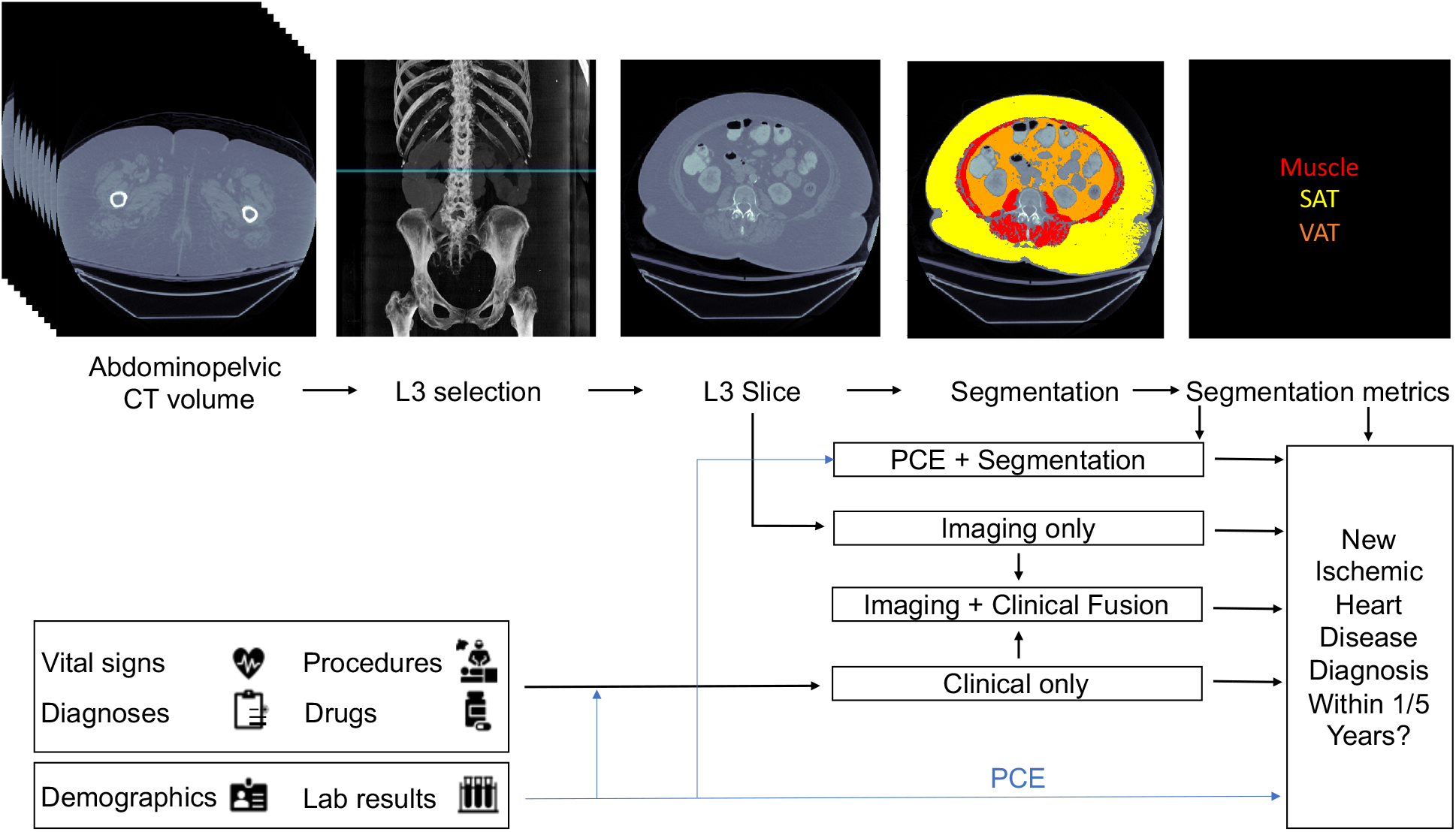
Proposed models for evaluating risk of a future ischemic heart disease event in one or five years following an abdominopelvic computed tomography (CT). The blue line shows which sources are used by the Pooled Cohort Equations (PCE), the standard tool for atherosclerotic cardiovascular disease risk assessment in current clinical guidelines for 10-year risk estimation. In our proposed models, the axial slice corresponding to the third lumbar vertebra anatomical level (L3) is automatically selected from the CT volume. In one model, the L3 slice is automatically segmented to extract mean muscle radiodensity in Hounsfield units and the Visceral/Adipose cross-sectional area ratio; these features are used alone or in combination with covariates from the PCE to form a segmentation or PCE + Segmentation model. Alternatively, features are automatically extracted from the L3 slice using a convolutional neural network (Imaging only model). As an additional approach, predictions from a model trained with features constructed from the patient’s electronic medical record within the year prior to CT acquisition (Clinical only model) are stacked with those of the imaging model (Imaging + Clinical fusion model).

**Figure 2.**
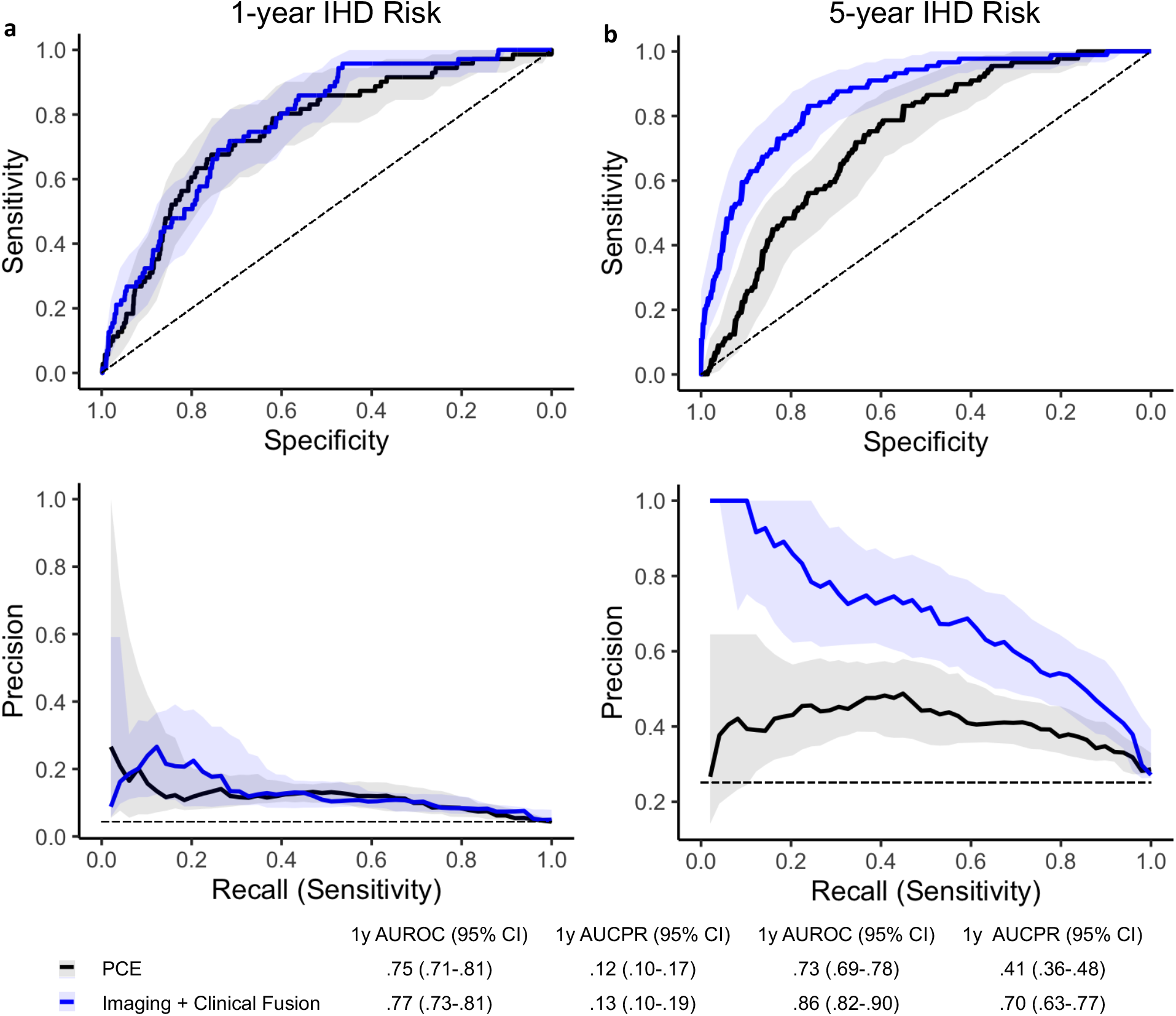
Performance of proposed Imaging + Clinical Fusion model compared to Pooled Cohort Equations (PCE), visualized through the receiver operating characteristic (ROC) curves (top row) and precision recall curves (bottom row) for (a) 1-year and (b) 5-year IHD risk modeling. Shaded lines indicate 95% confidence intervals (CI). Dashed lines show performance of a random (ROC curve) or a prevalence-based classifier (precision recall curve) as the simplest baselines. Area under the curve (AUC), 95% CI were determined using DeLong’s method for the ROC curve and using the bootstrap method for the precision-recall curve.

### Segmentation Model Performance

Example segmentations produced by the model are depicted in Figure 3a. These examples, along with a quantitative assessment of the model as measured by the Dice scores (Supplemental Figure 3), show that the model can reliably label the muscle and fat, with a mean (standard deviation) Dice score of .97 (.03), .97 (.02) and .96 (.05) for muscle, SAT and VAT, respectively. Furthermore, the error in computing tissue radiodensity and cross-sectional area was below 1% and 2%, respectively, for the three segmented tissues.

**Figure 3.**
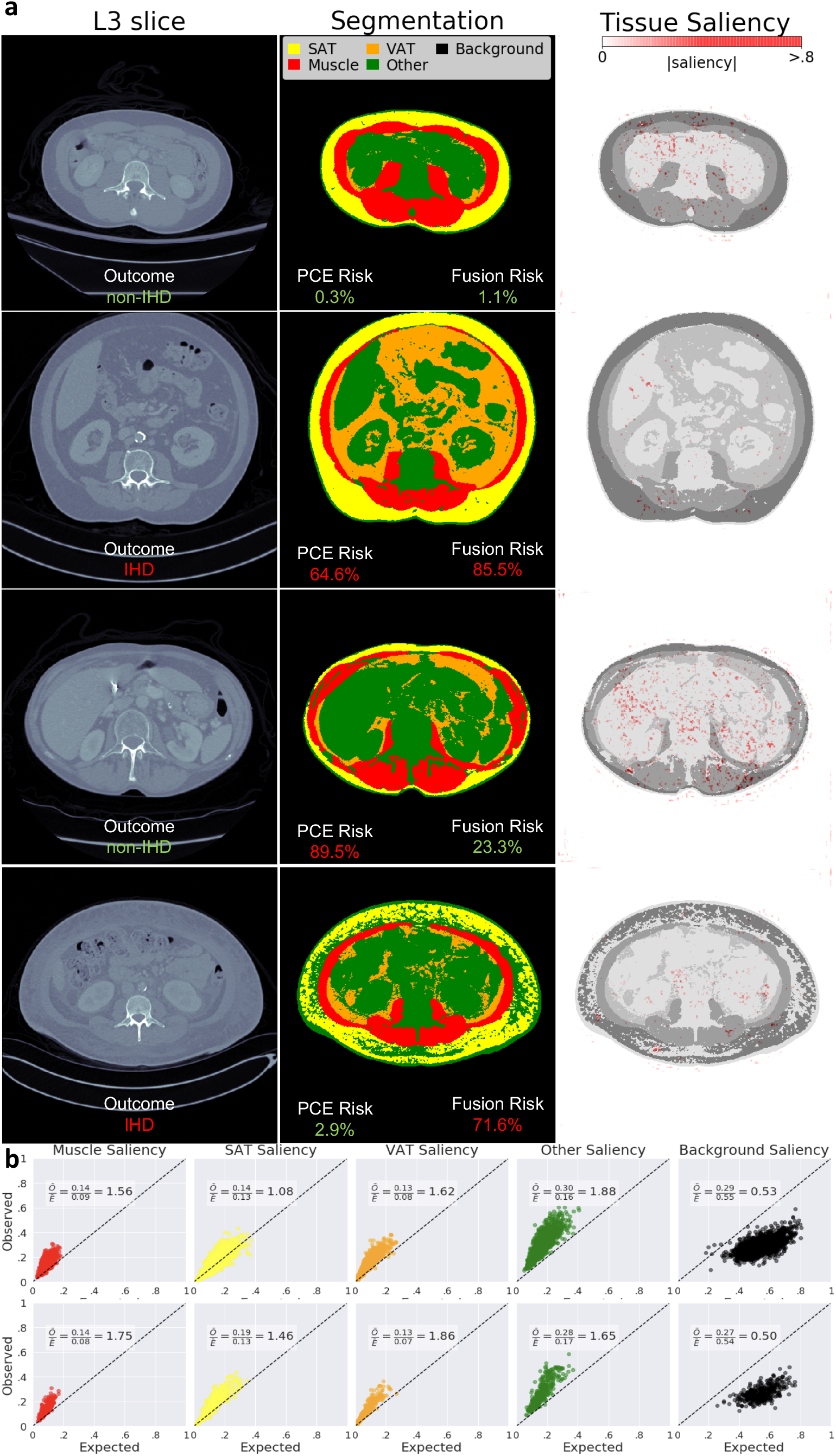
Segmentations and tissue saliency. In (a), sample L3 slices (first column) are shown for four individuals with at least 5-years follow-up after their image was acquired, with their corresponding segmentations generated from the segmentation model (second column). Their calculated risk from the traditional PCE is contrasted with the more accurate Imaging + Clinical fusion risk. The saliency from the imaging model is shown overlayed on the segmentation (third column). In (b) the distributions of observed (aggregated saliency values for each tissue type relative to the saliency for the image) versus expected saliency are shown for each tissue, for 1-year (top) and 5-year (bottom) risk prediction, where expected saliency is calculated as the proportion of pixels corresponding to a class in the image.

The bi-variate predictive model using VAT/SAT ratio and L3 muscle radiodensity as features (segmentation only model) performed comparably to the PCE, with 1-year AUROC of .70 (*P*=0.10) and AUCPR of .08 (*P*=.08) and 5-year AUROC of .73 (*P*=0.85) and AUCPR of .43 (*P*=.64).

### Imaging Only Model Performance

The Imaging Only Model also achieved comparable performance to the PCE for 1-year IHD risk prediction. It achieved a 1-year AUROC of .74 (*P*=.70) and AUCPR of .10 (*P*=.55). It showed improved performance in the 5-year risk prediction, both in terms of the AUROC (.81; *P*=.02), and AUCPR (.64; *P*<.001) (Table 1). This model also outperformed the Segmentation only model, with statistically significant increases of .04 (*P*=.74) and .09 (*P*=.01) in 1 and 5-year AUROC, respectively.

The contribution of individual tissues to the final prediction was assessed through tissue saliency. Figure 3a shows sample L3 slice segmentations, as well as tissue saliency values superimposed on the original image. Figure 3b shows the distribution of observed and expected tissue saliency values. For the 1-year follow-up cohort, the observed/expected tissue saliency ratios were 1.88, 1.62, 1.56, 1.08, and 0.53 for Other Tissues, VAT, muscle, SAT, and background respectively. For the 5-year follow-up cohort, the ratios were 1.86, 1.75, 1.65, 1.46, and 0.50 for VAT, muscle, Other Tissues, SAT, and background, respectively. That is, these ratios were higher than expected for muscle, VAT, SAT and other body tissues, and lower than expected for the background. All differences in pairs of observed vs. expected values were statistically significant (*P* < 0.001).

### Clinical Only Model Performance

The Clinical Only model achieved AUROC/AUCPR of .76/.12 and .84/.64 for 1 and 5-year IHD risk prediction, achieving comparable performance to the PCE in 1-year prediction (AUROC *P*=.57, AUCPR *P*=.98) and improved performance in 5-year prediction (*P*<.001 for AUROC and AUCPR).

Figure 4a shows the ten features with highest average SHAP value in the 5-year IHD risk prediction model. The same top 10 features were identified in the 1-year follow-up cohort, albeit not in the same order. In both cases, traditional cardiovascular risk factors, such as age, male gender and hypertension-related variables are present among the top features. The most impactful feature on prediction as determined by SHAP value was age, with higher risk for older individuals. SHAP values for 4 individuals from the 5-year risk cohort are shown in Figure 4b.

**Figure 4.**
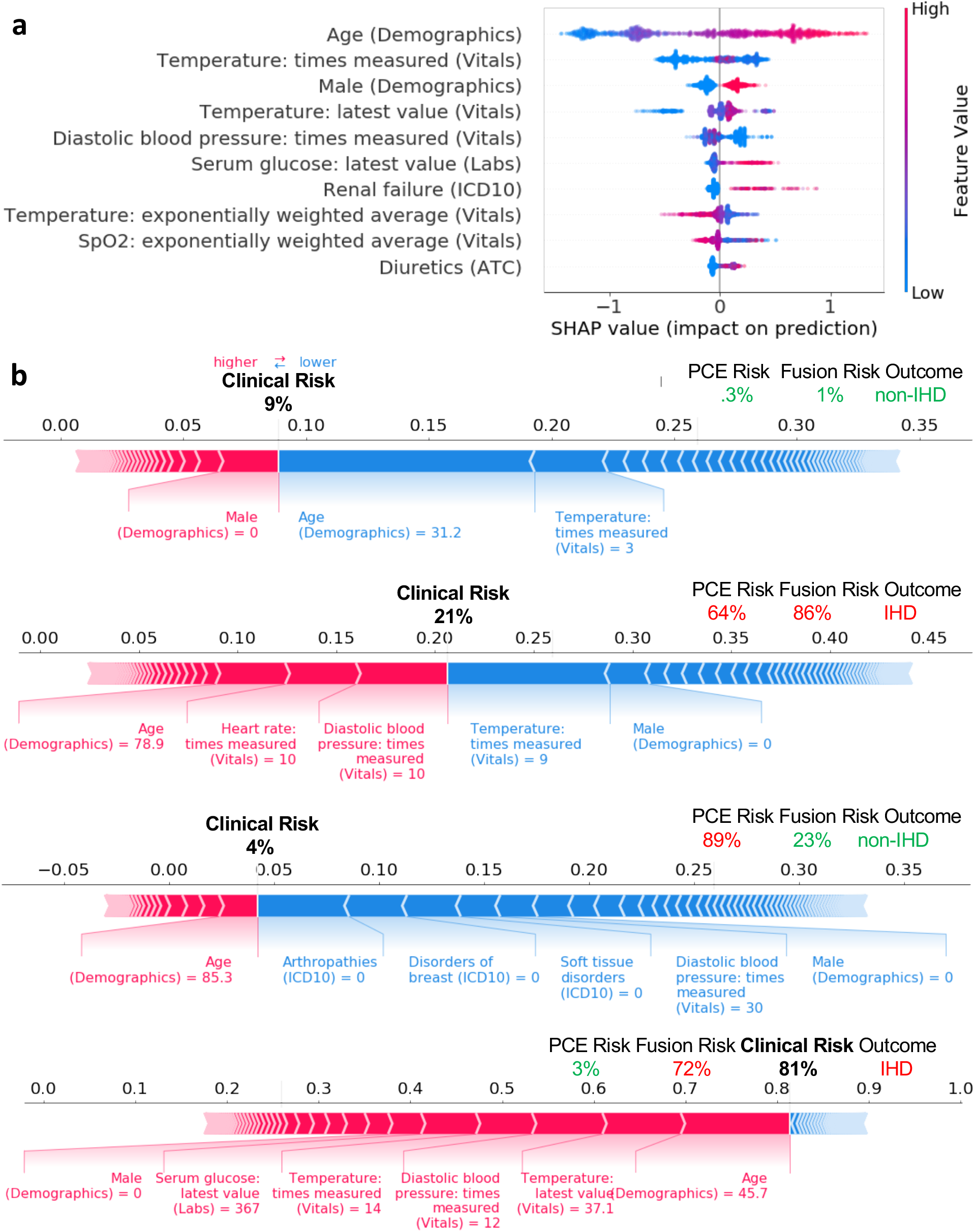
Clinical only model feature importance as quantified by SHAP (SHapley Additive exPlanations) values for the top 10 features in the training set of the 5-year risk prediction cohort (a). Higher SHAP values indicate higher than expected probability of IHD as assigned by the model. Individual SHAP values for features with highest values for 4 individuals from the 5-year risk prediction cohort (b), along with the risk assigned by the clinical only model. Their PCE and Imaging + Clinical Fusion model risk are also shown, along with the outcome.

### Fusion Model Performance

The performance of the predictive algorithms combining the BC segmentation metrics with the original PCE covariates (PCE + segmentation model) was comparable to that of the PCE alone, with 1 and 5-year AUROC/AUCPR values of .76 (*P*=.68)/.12 (*P*=.90) and .74 (*P*=.45)/.43 (*P*=.41), respectively. Thus, inclusion of average muscle radiodensity and VAT/SAT ratio did not improve prediction of IHD risk of PCE covariates.

The performance of the Imaging + Clinical fusion model is depicted in Figure 2. For 5-year IHD risk modeling, this model showed marked improvement in prediction capabilities compared to the PCE, both in terms of AUROC (.86) and AUCPR (.70), with a statistically significant increase (*P*<.001 in both metrics) of .12 and .29, respectively. For 1-year IHD risk modeling, the model performed comparably to the PCE, and there was no statistically significant difference.

The Imaging + Clinical + Segmentation fusion model performed similarly to the Imaging + Clinical fusion model. No statistically significant differences were found in AUROC or AUCPR for either 1 or 5-year IHD prediction when compared to the Imaging + Clinical fusion model. Furthermore, the same statistically significant increase in AUROC and AUCPR for 5-year IHD prediction compared to the PCE as the Imaging + Clinical fusion model was found.

The performance of baseline models and all proposed models across subpopulations with complete/missing PCE covariates, and across different age, gender, race/ethnicity, as well as those patients taking lipid-modifying agents is reported in Supplemental Table 5.

## Discussion

In this study we propose automated, explainable, and opportunistic risk assessment methods for 1-year and 5-year IHD risk following a contrast-enhanced abdominopelvic CT. Using a single slice from the CT, features quantifying the muscle radiodensity and body fat in conjunction with traditional cardiovascular risk factors and additional clinical data derived from the EMR, our models perform comparably or better than currently used tools to assess cardiovascular risk.

The use of BC biomarkers derived from abdominal CT imaging for cardiovascular risk assessment has been explored in the past. Pickhardt et al. extracted univariate metrics from CT colonography such as liver and muscle radiodensity, abdominal aortic calcification, and VAT/SAT ratio combined with FRS in asymptomatic individuals and found an improvement in 2-year cardiovascular event prediction AUROC of .77 compared to .71 using FRS alone^46^. While highlighting the value of imaging biomarkers in cardiovascular risk assessment, their methods were developed using CT colonography, an imaging modality that remains underutilized^47^. In contrast, our models perform opportunistic risk assessment in individuals that undergo contrast-enhanced abdominopelvic CTs, a more commonly used diagnostic scan in a wide variety of clinical settings. Thus, our model may potentially allow more opportunities for incidental risk assessment for IHD. Moreover, most diagnostic scans, such as abdominopelvic CTs, are geared towards answering a primary clinical question or indication for scan (e.g. cause for acute abdominal pain). Models such as the ones proposed in this study could increase the diagnostic and prognostic value of medical images by providing risk assessment in addition to the etiology of the patient’s acute symptoms.

Both the FRS and PCE have been shown to overestimate the risk of developing cardiovascular disease in contemporary, real-world populations^48^. As models used predominantly for time-to-event risk assessment, they have been developed to maximize the c-statistic (or AUROC in binary classification settings), and are typically used with cut-offs defined to have high sensitivity, at the expense of specificity. In our test cohorts, the AUROC of these baseline models was comparable to prior validation studies^4,49^. We believe that the use and reporting of AUCPR should be considered in the development of IHD risk assessment models, as it has been shown that 1) a curve will dominate in ROC if and only if it dominates in precision-recall space, and 2) PRC are more informative in an imbalanced classification setting, as is typical for IHD risk assessment^50^. By taking into account the trade-off between precision and recall, models can be designed to have a high sensitivity, but also have a high precision (i.e., fraction of true positives among those identified as positive). Our 5-year IHD risk models outperform the PCE in both the ROC and precision-recall space, which indicates that they can successfully identify individuals at risk of developing IHD, with a higher proportion of true positives among those identified with high risk.

Our methods seek to address model interpretability, an important barrier to implementing artificial intelligence in medicine^51^. Though our segmentation model had high Dice scores, similar to other published studies^52,53^, BC metrics alone or in combination with PCE covariates did not outperform the PCE. To our knowledge, our approach is the first to model IHD risk prediction in an end-to-end manner using the pixel data, as opposed to BC metrics, outperforming the radiomics/PCE approach in 5-year IHD prediction. In addition to treating IHD risk prediction as an end-to-end problem, we introduced the concept of tissue saliency to study the contribution of pixels to the predictions made by the Imaging only Model. This enabled us to assess the contribution of groups of pixels belonging to the same tissue class in a qualitative and quantitative manner. As expected, tissues within the body had a higher amount of saliency than expected, most notably the VAT and muscle tissues. This is consistent with the observation that biomarkers quantifying the radiodensity or area of these tissues are informative of IHD risk^15^. The tissue saliency ratio for *Other Tissues* was also higher than expected, which could be due to the liver, abdominal aortic calcifications or trabecular bone radiodensity being present in slices at this level, but not explicitly segmented in this study (as can be seen in examples of Figure 3a). Furthermore, the background pixels provided a lower-than-expected but not negligible proportion of the saliency. Upon visualization of examples, background pixels with higher saliency are typically neighboring the body, which indicates that the model may be using patient habitus as a feature. In aggregate, tissue saliency provides insights into the contribution of tissues in the prediction, increasing our understanding of the underlying drivers of prediction in the better-performing Imaging only Model.

We found evidence that including additional clinical features could improve the performance of the Imaging only Model. We examined the importance of individual clinical features through their SHAP values. Among the top predictors were features that represent traditional cardiovascular risk factors, such as age, male gender, and hypertension. Features such as increased serum glucose, which is associated with a higher risk in our models, may better assess IHD risk than the diagnosis of diabetes alone, a feature which was not salient among the SHAP analysis. Similarly, the use of diuretics -a common treatment for hypertension-was positively associated with IHD; hypertension is a well-studied modifiable IHD risk factor^54^. Finally, renal failure, the only diagnosis code among the top ten predictors, has also been identified as an independent risk factor for IHD^55^. The use of SHAP values aids in understanding how an individual feature may affect the prediction of IHD risk. The presence of well-known risk factors among the top predictors raises trust in an individual attempting to scrutinize the prediction model. SHAP values and tissue saliency could provide clinicians with a mechanism to interpret and intervene based on specific aspects of the patient history. As new artificial intelligence applications in medical imaging continue to emerge and gain popularity, explainability may be an important key factor for clinical adoption^56,57^.

There are several potential ways our models could be utilized in clinical practice. After undergoing a commonly performed contrast-enhanced abdominopelvic CT for non-IHD indications, individuals could be opportunistically assessed for high IHD risk and undergo further cardiovascular follow-up or be referred to primary care or cardiology for potential initiation of preventive strategies. Furthermore, the specific BC metrics automatically calculated from the CT could be used to identify tangible areas of improvement, as well as be used to track progress following an intervention. Ultimately, these models could identify an individual at high IHD risk that may have gone otherwise unnoticed, which is the goal of opportunistic risk assessment. Finally, the models could analyze CT scans that have already been performed and are housed within a picture archiving and communication system for retrospective identification of high IHD risk patients.

While promising, our study has limitations. Our data were sourced retrospectively from a single center. Though our reported results correspond to a test set of patients that was not used during model development, the confirmation of the potential clinical use of the models would require a prospective evaluation, ideally in multiple centers. Such evaluations could also address a potential selection bias in our study, where we selected patients that present to the emergency department with abdominal pain and have an contrast-enhanced abdominopelvic CT scan performed. In addition, though our cohorts were comprised of diverse individuals both in terms of gender and ethnicity (Supplemental Table 3), these have been identified as variables across which model performance may vary, typically to the disadvantage of underrepresented minorities^58^. We found small variations to be present within patient subpopulations (Supplemental Table 5), motivating further studies in other validation cohorts to identify demographic-specific thresholds for intervention^59^. Another limitation is that biomarkers such as aortic calcifications or liver radiodensity may not necessarily be visualized in the single L3 slice approach that we analyzed. This may explain the lack of improvement when combining segmentation metrics with PCE features, that has been identified in other studies^17^. Finally, our 5-year cohort prediction models showed an improvement over established baselines, but our 1-year IHD risk prediction models were not able to outperform them, despite exploring a variety of data sampling and alternative loss functions to improve the performance of these models. This improved performance in 5-year prediction, however, could provide a window of opportunity to initiate preventive interventions. Alternatively, a considerable proportion of individuals in our cohort were missing laboratory measurements that would be necessary to evaluate the PCE or FRS at the time of scan acquisition, indicating their cardiovascular risk was not recently assessed. Our opportunistic approach could be used to alert a referring provider of particularly high IHD risk individuals and prompt further evaluation.

In conclusion, we develop a framework to use artificial intelligence models that enable opportunistic risk assessment for IHD following an abdominopelvic CT scan. By drawing from multiple data sources, we were able to produce models that can perform comparably or better than currently used clinical risk models, which currently guide treatment decisions for individuals undergoing cardiovascular risk assessment. Models automatically integrating existing EMR and CT data could provide opportunities for more effective preventive IHD interventions at a population health level.

## Code availability

In order to facilitate replication of these results and further research, we will make our code and trained segmentation model publicly available at http://github.com/ChaudhariLab/AbCT_IHD.

## Data availability

Our model predictions and outcome labels will be made publicly available alongside our code. The raw data are not publicly available due to privacy information embedded directly within the data. Data are available on request due to privacy or other restrictions. The data that support the findings of this study are available on request from the corresponding author (B.P.).

## Author Contributions

JMZC performed all experiments and wrote the manuscript. ASC helped with experiments, writing the manuscript, creating figures, and manuscript editing. AD developed the segmentation model. AW performed segmentations and provided manuscript edits. RB, DM, FR, AS, BJ, and DR provided manuscript edits. BP designed the concept and study, oversaw the experiments, and edited the manuscript.

