## Supplemental Figures and Tables for "Opportunistic Assessment of Ischemic Heart Disease Risk Using Abdominopelvic Computed Tomography and Medical Record Data: a Multimodal Explainable Artificial Intelligence Approach"

### Appendix: Supplemental Figures and Tables

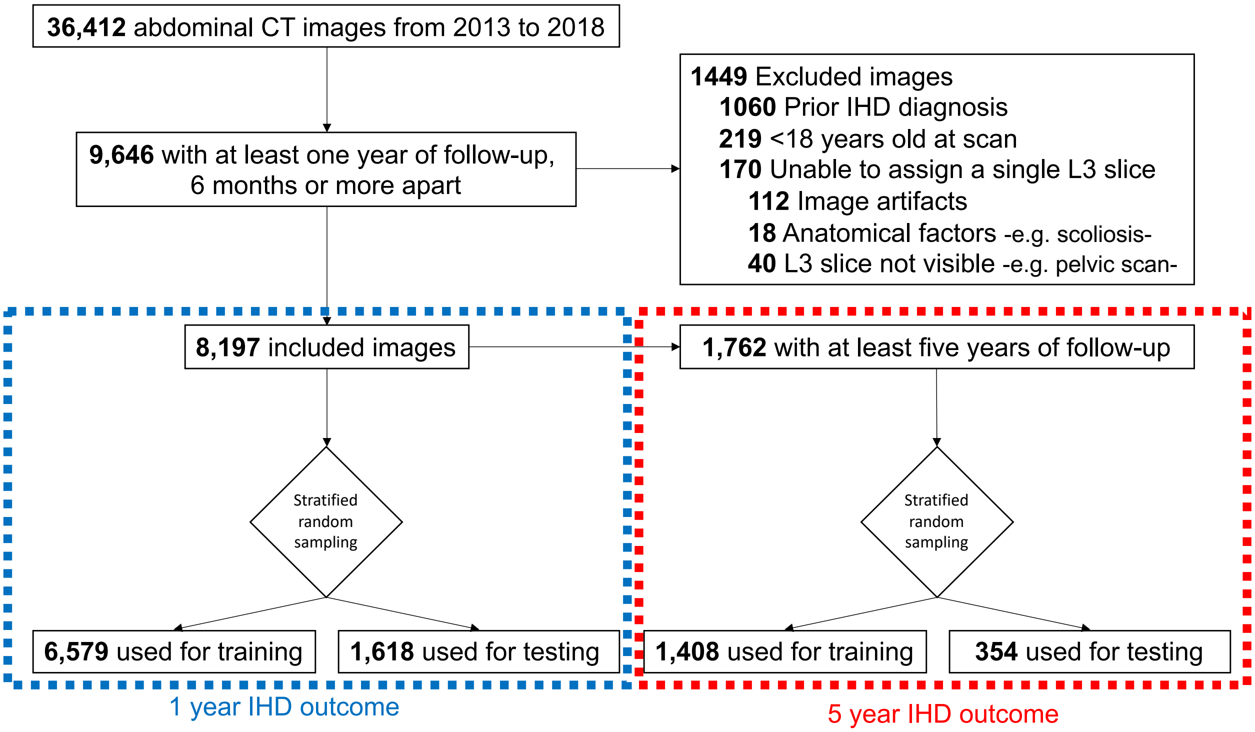


Supplemental Figure 1. Sampling criteria to arrive at final cohorts.

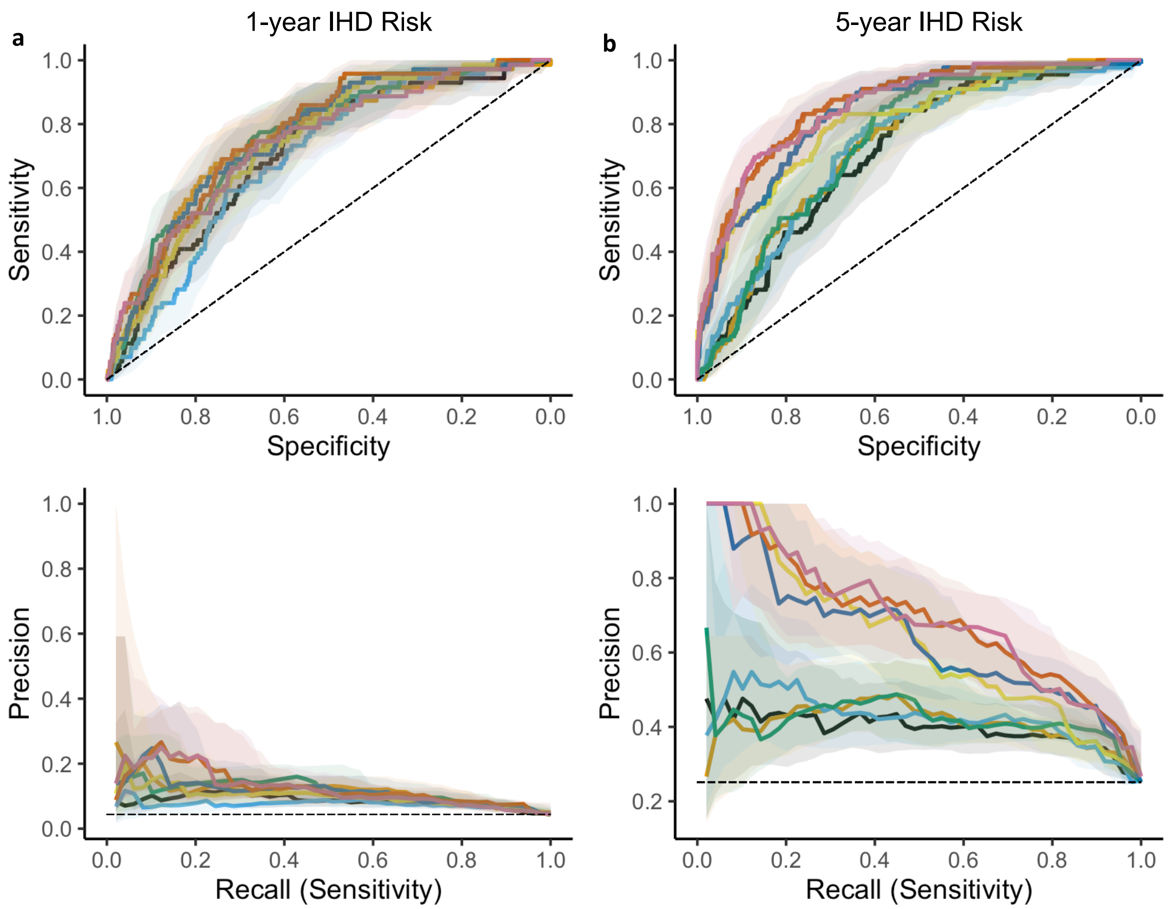


| | 1y AUROC (95% CI) | 1y AUCPR (95% CI) | 1y AUROC (95% CI) | 1y AUCPR (95% CI) |
|---|---|---|---|---|
| FRS | .71 (.67-.76) | .09 (.07-.12) | .71 (.66-.76) | .40 (.35-.48) |
| PCE | .75 (.71-.81) | .12 (.10-.17) | .73 (.69-.78) | .41 (.36-.48) |
| Segmentation only (S) | .70 (.65-.74) | .08 (.07-.10) | .73 (.68-.78) | .43 (.38-.51) |
| PCE + Segmentation (PCE + S) | .76 (.71-.81) | .12 (.10-.15) | .74 (.70-.79) | .43 (.38-.51) |
| Imaging only (I) | .76 (.72-.81) | .12 (.10-.17) | .84 (.80-.87) | .64 (.58-.72) |
| Clinical Only (C) | .74 (.70-.78) | .10 (.08-.15) | .81 (.76-.85) | .64 (.57-.71) |
| Imaging + Clinical Fusion (I+C) | **.77 (.73-.81)** | **.13 (.10-.19)** | **.86 (.82-.90)** | **.70 (.63-.77)** |
| Imaging + Clinical + Segmentation Fusion (I+C+S) | .74 (.70-.79) | **.13 (.10-.18)** | **.86 (.82-.89)** | **.70 (.63-.77)** |

1y AUROC *P*-value

| Model | FRS | PCE | S | PCE+S | C | I | I+C | I+C+S |
|---|---|---|---|---|---|---|---|---|
| FRS | - | **.04** | .60 | **.007** | .05 | .39 | **.03** | .37 |
| PCE | - | - | .10 | .68 | .57 | .70 | .38 | .74 |
| S | - | - | - | **.02** | **.03** | .14 | **.009** | .22 |
| PCE+S | - | - | - | - | .79 | .53 | .56 | .57 |
| C | - | - | - | - | - | .37 | .55 | .25 |
| I | - | - | - | - | - | - | **.03** | .91 |
| I+C | - | - | - | - | - | - | - | **.010** |
| I+C+S | - | - | - | - | - | - | - | - |

5y AUROC *P*-value

| Model | FRS | PCE | S | PCE+S | C | I | I+C | I+C+S |
|---|---|---|---|---|---|---|---|---|
| FRS | - | .24 | .55 | .11 | **<.001** | **.005** | **<.001** | **<.001** |
| PCE | - | - | .85 | .45 | **<.001** | **.02** | **<.001** | **<.001** |
| S | - | - | - | .54 | **<.001** | **.01** | **<.001** | **<.001** |
| PCE+S | - | - | - | - | **<.001** | **.04** | **<.001** | **<.001** |
| C | - | - | - | - | - | .29 | **.03** | .14 |
| I | - | - | - | - | - | - | **.003** | **.01** |
| I+C | - | - | - | - | - | - | - | .41 |
| I+C+S | - | - | - | - | - | - | - | - |

1y AUCPR *P*-value

| Model | FRS | PCE | S | PCE+S | C | I | I+C | I+C+S |
|---|---|---|---|---|---|---|---|---|
| FRS | - | .06 | .46 | **.04** | .15 | .51 | .09 | .14 |
| PCE | - | - | .08 | .90 | .98 | .55 | .73 | .78 |
| S | - | - | - | **.02** | .08 | .19 | **.04** | .06 |
| PCE+S | - | - | - | - | .93 | .54 | .59 | .67 |
| C | - | - | - | - | - | .53 | .47 | .56 |
| I | - | - | - | - | - | - | .15 | .21 |
| I+C | - | - | - | - | - | - | - | .77 |
| I+C+S | - | - | - | - | - | - | - | - |

5y AUCPR *P*-values

| Model | FRS | PCE | S | PCE+S | C | I | I+C | I+C+S |
|---|---|---|---|---|---|---|---|---|
| FRS | - | .73 | .54 | .38 | **<.001** | **<.001** | **<.001** | **<.001** |
| PCE | - | - | .64 | .41 | **<.001** | **<.001** | **<.001** | **<.001** |
| S | - | - | - | .93 | **<.001** | **<.001** | **<.001** | **<.001** |
| PCE+S | - | - | - | - | **<.001** | **<.001** | **<.001** | **<.001** |
| C | - | - | - | - | - | .94 | **.007** | **.01** |
| I | - | - | - | - | - | - | .14 | .15 |
| I+C | - | - | - | - | - | - | - | .99 |
| I+C+S | - | - | - | - | - | - | - | - |

Supplemental Figure 2. Proposed model performance in comparison to pooled cohort Equations (PCE) and Framingham risk score (FRS) as measured by receiver operating characteristic and precision-recall curves for 1-year (a) and 5-year (b) IHD risk prediction. Shaded areas correspond to 95% confidence intervals. In figure legend area under receiver operating characteristic (AUROC) and precision-recall (AUCPR) curves are shown. Below, *P* values for pair-wise comparisons between models are reported. Confidence intervals (CI) and *P* values were obtained using the DeLong method for AUROC and the bootstrap method for AUCPR.

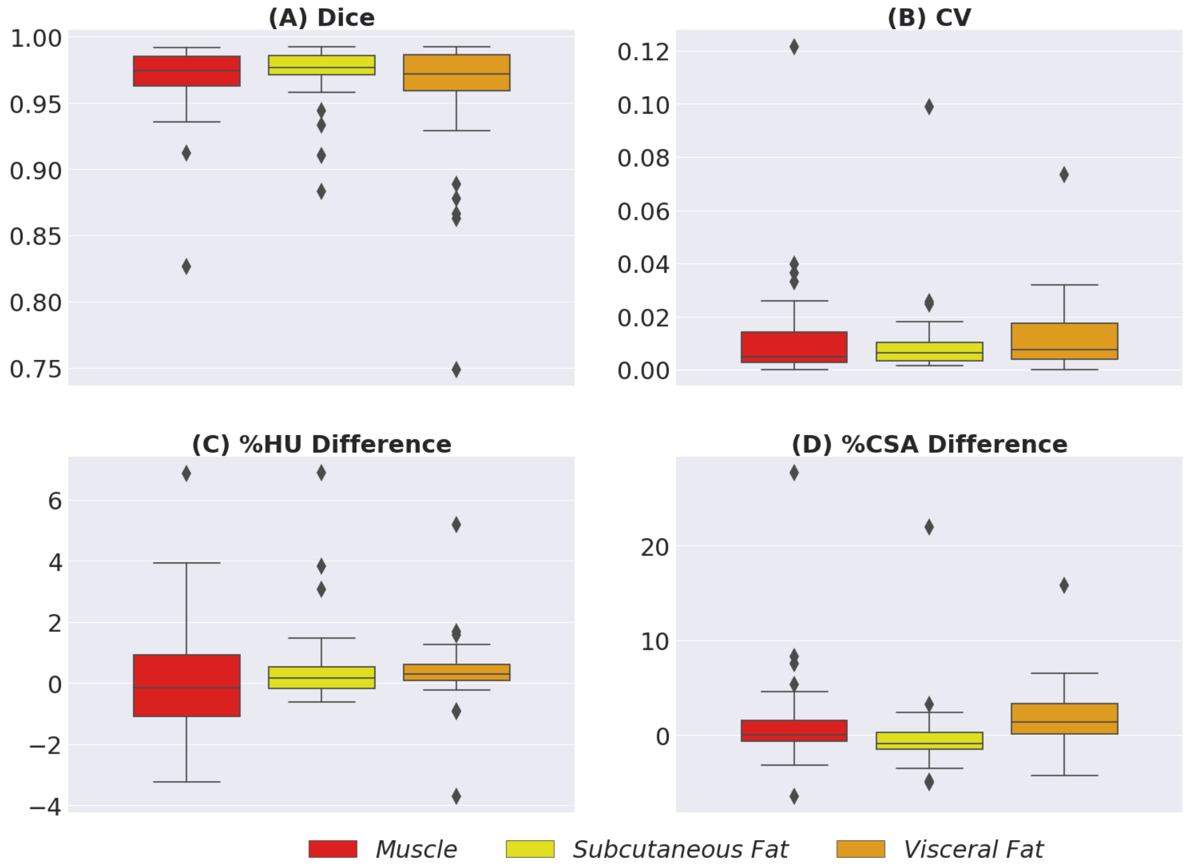


Supplemental Figure 3. Segmentation model performance. CV : coefficient of variation, HU: Hounsfield Unit, CSA: cross-sectional area. Center line represents median, box limits upper and lower quartiles, whiskers 1.5x interquartile range, and points outliers.

| Model | Hyperparameters tested | Best Validation AUROC | |
|---|---|---|---|
| | | 1y | 5y |
| ***Segmentation only*** | *C: {.001, 0.1, 0.5, 1.0,5**,10}* | *.6757* | *.6907* |
| ***Imaging only*** | | | |
| *Selected configuration* | *Batch size 8*<br>*Learning rate {1e-5*,1e-6**}*<br>*Adam optimizer*<br>*Max epochs {10*,30**}*<br>*Cross Entropy Loss* | *.7331* | *.7969* |
| *Training only final layer* | *Batch size 8*<br>*Learning rate {1e-4,1e-5,1e-6}*<br>*Adam optimizer*<br>*Max epochs 15*<br>*Cross Entropy Loss* | *.5656* | *.6523* |
| *Image augmentations* | *Batch size 8*<br>*Learning rate {1e-3, 1e-4, 1e-5, 1e-6}*<br>*Adam optimizer*<br>*Max Epochs 20*<br>*Cross Entropy Loss*<br>*Rotations of up to 3° and pixel shifting of up to 5 pixels during training* | *.7089* | *.7867* |
| *Focal loss* | *Batch size 8*<br>*Learning rate {1e-4,1e-5, 1e-6}*<br>*Adam optimizer*<br>*Max epochs 20*<br>*Focal loss (gamma:{2,3,4,5,10} alpha:{[.01,.99],[0.05,0.95]})*<br>*Augmentations {Yes/No}* | *.7275* | *.7879* |
| ***Clinical only*** | *colsample_bytree:{.2,.4**,.6,.8,1*}*<br>*gamma:{.1,.5**,1,10*}*<br>*max_depth: {1,2,3,4,5*,**,6}*<br>*min_child_weight: {.1,.5**,1*}*<br>*subsample: {.5*, .75**, 1}*<br>*scale_pos_weight:{1*,**,5,10,15,20}* | *.7851* | *.7935* |
| ***PCE + segmentation*** | *colsample_bytree:{.2*,**,.4,.6,.8,1}*<br>*gamma:{.1,.5,1,10*,**}*<br>*max_depth: {1,2,3*,4,5**,6}*<br>*min_child_weight: {.1*,.5,1**}*<br>*subsample: {.5**, .75, 1*}* | *.7427* | *.7262* |
| ***Imaging + Clinical Fusion*** | *C: {0.1, 0.5, 1.0,5**,10,20,30, 50, 100*, 500, 1000}* | *.9737* | *.9751* |
| ***Imaging + Clinical + Segmentation Fusion*** | *C: {0.1, 0.5, 1.0,5**,10,20,30, 50, 100*, 500, 1000*,**}* | *.9777* | *.9805* |

Supplemental Table 1. Validation performance of alternative training schemes and hyperparameters of predictive models. *,**: hyperparameters selected for 1-year and 5-year IHD prediction, respectively.

Supplemental Table 2. Variables captured from the electronic medical record and used as covariates of Clinical only model. (See attached file).

| Variable | 1-year follow-up | | | | | | | | 5-year follow-up | | | | | | | |
|---|---|---|---|---|---|---|---|---|---|---|---|---|---|---|---|---|
| | Train | | | | Test | | | | Train | | | | Test | | | |
| | non-IHD (n=6292) | | IHD (n=287) | | non-IHD (n=1547) | | IHD (n=71) | | non-IHD (n=1051) | | IHD (n=357) | | non-IHD (n=265) | | IHD (n=89) | |
| | % / Mean (SD) | missing | % / Mean (SD) | missing | % / Mean (SD) | missing | % / Mean (SD) | missing | % / Mean (SD) | missing | % / Mean (SD) | missing | % / Mean (SD) | missing | % / Mean (SD) | missing |
| Age | 51.2 (17.1) | 0 | 63.9 (14.3) | 0 | 50.9 (16.5) | 0 | 65.8 (15.4) | 0 | 51.6 (16.4) | 0 | 63.9 (15.3) | 0 | 50.6 (16.9) | 0 | 62.7 (13.8) | 0 |
| Male Sex | 39.5 | 0 | 51.2 | 0 | 41.8 | 0 | 52.1 | 0 | 37.6 | 0 | 51.5 | 0 | 32.8 | 0 | 42.7 | 0 |
| Smoking | 6.2 | 177 | 6.3 | 1 | 6.6 | 59 | 1.45 | 2 | 5.7 | 32 | 7.1 | 3 | 5.0 | 7 | 3.4 | 0 |
| Race/Ethnicity | | 0 | | 0 | | 0 | | 0 | | 0 | | 0 | | 0 | | 0 |
| Non-Hispanic White | 48.0 | | 43.2 | | 49.5 | | 50.7 | | 52.9 | | 45.6 | | 51.4 | | 41.7 | |
| Non-Hispanic Black | 3.9 | | 2.8 | | 4.0 | | 2.8 | | 3.1 | | 2.4 | | 3.6 | | 4.2 | |
| Hispanic | 21.5 | | 20.9 | | 18.8 | | 21.1 | | 17.8 | | 20.0 | | 21.3 | | 26.4 | |
| Asian | 16.2 | | 20.9 | | 15.9 | | 15.5 | | 15.1 | | 19.6 | | 14.5 | | 19.4 | |
| Other/Unknown | 10.3 | | 12.2 | | 11.8 | | 9.9 | | 11.1 | | 12.6 | | 9.2 | | 8.3 | |
| Treated for hypertension | 25.4 | 0 | 49.5 | 0 | 23.6 | 0 | 52.1 | 0 | 28.4 | 0 | 51.0 | 0 | 25.7 | 0 | 49.4 | 0 |
| Diabetes diagnosis | 13.9 | 0 | 28.2 | 0 | 12.9 | 0 | 16.9 | 0 | 12.4 | 0 | 25.8 | 0 | 13.6 | 0 | 27.0 | 0 |
| HDL Cholesterol [mg/dL] | 56.9 (20.3) | 5113 | 57.3 (22.7) | 226 | 56.4 (20.2) | 1244 | 57.6 (14.2) | 57 | 56.9 (23.4) | 840 | 55.8 (19.5) | 269 | 55.3 (17.9) | 211 | 61.8 (24.2) | 69 |
| Total Cholesterol [mg/dL] | 183.1 (45.7) | 5039 | 173.9 (48.1) | 224 | 189.0 (47.5) | 1238 | 178.4 (49.4) | 53 | 187.4 (45.4) | 825 | 177.2 (46.9) | 262 | 177.7 (40.9) | 209 | 184.7 (41.4) | 69 |
| Systolic Blood Pressure [mmHg] | 124.3 (18.0) | 124 | 129.7 (20.9) | 8 | 124.2 (17.9) | 25 | 128.8 (18.5) | 4 | 123.6 (18.1) | 33 | 128.7 (21) | 8 | 123.8 (17.1) | 6 | 131.3 (20.4) | 6 |
| VAT/SAT ratio | .68 (1.6) | 0 | .86 (.7) | 0 | .66 (.7) | 0 | .85 (.6) | 0 | .63 (.6) | 0 | .87 (.7) | 0 | .62 (.5) | 0 | .80 (.6) | 0 |
| Muscle density [HU] | 43.9 (9.6) | 0 | 38.2 (9.1) | 0 | 43.7 (9.6) | 0 | 37.7 (7.9) | 0 | 44.0 (9.1) | 0 | 38.7 (8.9) | 0 | 44.3 (9.2) | 0 | 37.4 (8.8) | 0 |

Supplemental Table 3. Distribution of demographic variables across the study population. Features relevant to pooled cohort equations risk assessment, as well as body composition metrics derived from segmentation are also included.

| Model | 1-year IHD risk | | | | 5-year IHD risk | | | |
|---|---|---|---|---|---|---|---|---|
| | Sensitivity [%] | Specificity [%] | PPV [%] | NPV [%] | Sensitivity [%] | Specificity [%] | PPV [%] | NPV [%] |
| PCE ( $\geq$ 7.5% threshold) | 67.6 | 72.3 | 10.1 | 98.0 | 56.2 | 75.5 | 10.1 | 83.7 |
| PCE ( $\geq$ 20% threshold) | 35.2 | 87.8 | 11.7 | 96.7 | 31.5 | 87.5 | 11.7 | 79.2 |
| Segmentation only | 80.3 | 53.0 | 7.3 | 98.3 | 78.7 | 62.6 | 41.4 | 89.7 |
| Clinical Only | 69.0 | 71.6 | 10.0 | 98.1 | 84.3 | 70.6 | 49.0 | 93.0 |
| Imaging Only | 70.4 | 66.8 | 8.9 | 98.0 | 78.7 | 71.3 | 47.9 | 90.9 |
| Imaging + Clinical Fusion | 71.8 | 71.6 | 10.4 | 98.2 | 83.1 | 76.2 | 54.0 | 93.1 |

Supplemental Table 4. Performance of proposed classification models using Youden's index as threshold, compared to two clinically relevant thresholds for pooled cohort equations (PCE).

Supplemental Table 5. Performance of proposed prediction models, along with Framingham Risk Score (FRS) and Pooled Cohort Equations (PCE) in different subpopulations of the test set. y/o: years old. (See attached file).
