## Supplemental Table 2 for "Opportunistic Assessment of Ischemic Heart Disease Risk Using Abdominopelvic Computed Tomography and Medical Record Data: a Multimodal Explainable Artificial Intelligence Approach"

| Variable | Category | Name | Description |
| --- | --- | --- | --- |
| A01 | ATC | Stomatological Preparations | Number of prescriptions that match Anatomical Therapeutic Chemical code classification, 2nd level: A01 |
| A02 | ATC | Drugs For Acid Related Disorders | Number of prescriptions that match Anatomical Therapeutic Chemical code classification, 2nd level: A02 |
| A03 | ATC | Drugs For Functional Gastrointestinal Disorders | Number of prescriptions that match Anatomical Therapeutic Chemical code classification, 2nd level: A03 |
| A04 | ATC | Antiemetics And Antinauseants | Number of prescriptions that match Anatomical Therapeutic Chemical code classification, 2nd level: A04 |
| A05 | ATC | Bile And Liver Therapy Drugs | Number of prescriptions that match Anatomical Therapeutic Chemical code classification, 2nd level: A05 |
| A06 | ATC | Drugs For Constipation | Number of prescriptions that match Anatomical Therapeutic Chemical code classification, 2nd level: A06 |
| A07 | ATC | Antidiarrheals, Intestinal Antiinflammatory/Antiinfec tive Agents | Number of prescriptions that match Anatomical Therapeutic Chemical code classification, 2nd level: A07 |
| A08 | ATC | Antiobesity Preparations, Excl. Diet Products | Number of prescriptions that match Anatomical Therapeutic Chemical code classification, 2nd level: A08 |
| A09 | ATC | Digestives, Incl. Enzymes | Number of prescriptions that match Anatomical Therapeutic Chemical code classification, 2nd level: A09 |
| A10 | ATC | Drugs Used In Diabetes | Number of prescriptions that match Anatomical Therapeutic Chemical code classification, 2nd level: A10 |
| A11 | ATC | Vitamins | Number of prescriptions that match Anatomical Therapeutic Chemical code classification, 2nd level: A11 |

| | | | |
|---|---|---|---|
| A12 | ATC | Mineral Supplements | Number of prescriptions that match Anatomical Therapeutic Chemical code classification, 2nd level: A12 |
| A14 | ATC | Anabolic Agents For Systemic Use | Number of prescriptions that match Anatomical Therapeutic Chemical code classification, 2nd level: A14 |
| A16 | ATC | Other Alimentary Tract And Metabolism Products In Atc | Number of prescriptions that match Anatomical Therapeutic Chemical code classification, 2nd level: A16 |
| B01 | ATC | Antithrombotic Agents | Number of prescriptions that match Anatomical Therapeutic Chemical code classification, 2nd level: B01 |
| B02 | ATC | Antihemorrhagics | Number of prescriptions that match Anatomical Therapeutic Chemical code classification, 2nd level: B02 |
| B03 | ATC | Antianemic Preparations | Number of prescriptions that match Anatomical Therapeutic Chemical code classification, 2nd level: B03 |
| B05 | ATC | Blood Substitutes And Perfusion Solutions | Number of prescriptions that match Anatomical Therapeutic Chemical code classification, 2nd level: B05 |
| B06 | ATC | Other Hematological Agents In Atc | Number of prescriptions that match Anatomical Therapeutic Chemical code classification, 2nd level: B06 |
| C01 | ATC | Cardiac Therapy Drugs | Number of prescriptions that match Anatomical Therapeutic Chemical code classification, 2nd level: C01 |
| C02 | ATC | Antihypertensives | Number of prescriptions that match Anatomical Therapeutic Chemical code classification, 2nd level: C02 |
| C03 | ATC | Diuretics | Number of prescriptions that match Anatomical Therapeutic Chemical code classification, 2nd level: C03 |
| C04 | ATC | Peripheral Vasodilators | Number of prescriptions that match Anatomical Therapeutic Chemical code classification, 2nd level: C04 |

| | | | |
|---|---|---|---|
| C05 | ATC | Vasoprotectives | Number of prescriptions that match Anatomical Therapeutic Chemical code classification, 2nd level: C05 |
| C07 | ATC | Beta-Adrenergic Blocking Agents | Number of prescriptions that match Anatomical Therapeutic Chemical code classification, 2nd level: C07 |
| C08 | ATC | Calcium Channel Blockers | Number of prescriptions that match Anatomical Therapeutic Chemical code classification, 2nd level: C08 |
| C09 | ATC | Agents Acting On The Renin-Angiotensin System | Number of prescriptions that match Anatomical Therapeutic Chemical code classification, 2nd level: C09 |
| C10 | ATC | Lipid Modifying Agents | Number of prescriptions that match Anatomical Therapeutic Chemical code classification, 2nd level: C10 |
| D01 | ATC | Antifungals For Dermatological Use | Number of prescriptions that match Anatomical Therapeutic Chemical code classification, 2nd level: D01 |
| D02 | ATC | Emollients And Protectives | Number of prescriptions that match Anatomical Therapeutic Chemical code classification, 2nd level: D02 |
| D03 | ATC | Preparations For Treatment Of Wounds And Ulcers | Number of prescriptions that match Anatomical Therapeutic Chemical code classification, 2nd level: D03 |
| D04 | ATC | Antipruritics, Incl. Antihistamines, Anesthetics, Etc. | Number of prescriptions that match Anatomical Therapeutic Chemical code classification, 2nd level: D04 |
| D05 | ATC | Antipsoriatics | Number of prescriptions that match Anatomical Therapeutic Chemical code classification, 2nd level: D05 |
| D06 | ATC | Antibiotics And Chemotherapeutics For Dermatological Use | Number of prescriptions that match Anatomical Therapeutic Chemical code classification, 2nd level: D06 |
| D07 | ATC | Corticosteroids, Dermatological Preparations | Number of prescriptions that match Anatomical Therapeutic Chemical code classification, 2nd level: D07 |

| | | | |
|---|---|---|---|
| D08 | ATC | Antiseptics And Disinfectants | Number of prescriptions that match Anatomical Therapeutic Chemical code classification, 2nd level: D08 |
| D09 | ATC | Medicated Dressings | Number of prescriptions that match Anatomical Therapeutic Chemical code classification, 2nd level: D09 |
| D10 | ATC | Anti-Acne Preparations | Number of prescriptions that match Anatomical Therapeutic Chemical code classification, 2nd level: D10 |
| D11 | ATC | Other Dermatological Preparations In Atc | Number of prescriptions that match Anatomical Therapeutic Chemical code classification, 2nd level: D11 |
| G01 | ATC | Gynecological Antiinfectives And Antiseptics | Number of prescriptions that match Anatomical Therapeutic Chemical code classification, 2nd level: G01 |
| G02 | ATC | Other Gynecologicals In Atc | Number of prescriptions that match Anatomical Therapeutic Chemical code classification, 2nd level: G02 |
| G03 | ATC | Sex Hormones And Modulators Of The Genital System | Number of prescriptions that match Anatomical Therapeutic Chemical code classification, 2nd level: G03 |
| G04 | ATC | Urologicals | Number of prescriptions that match Anatomical Therapeutic Chemical code classification, 2nd level: G04 |
| H01 | ATC | Pituitary And Hypothalamic Hormones And Analogues | Number of prescriptions that match Anatomical Therapeutic Chemical code classification, 2nd level: H01 |
| H02 | ATC | Corticosteroids For Systemic Use | Number of prescriptions that match Anatomical Therapeutic Chemical code classification, 2nd level: H02 |
| H03 | ATC | Thyroid Therapy Drugs | Number of prescriptions that match Anatomical Therapeutic Chemical code classification, 2nd level: H03 |
| H04 | ATC | Pancreatic Hormones | Number of prescriptions that match Anatomical Therapeutic Chemical code classification, 2nd level: H04 |

| | | | |
|---|---|---|---|
| H05 | ATC | Calcium Homeostasis | Number of prescriptions that match Anatomical Therapeutic Chemical code classification, 2nd level: H05 |
| J01 | ATC | Antibacterials For Systemic Use | Number of prescriptions that match Anatomical Therapeutic Chemical code classification, 2nd level: J01 |
| J02 | ATC | Antimycotics For Systemic Use | Number of prescriptions that match Anatomical Therapeutic Chemical code classification, 2nd level: J02 |
| J04 | ATC | Antimycobacterials | Number of prescriptions that match Anatomical Therapeutic Chemical code classification, 2nd level: J04 |
| J05 | ATC | Antivirals For Systemic Use | Number of prescriptions that match Anatomical Therapeutic Chemical code classification, 2nd level: J05 |
| J06 | ATC | Immune Sera And Immunoglobulins | Number of prescriptions that match Anatomical Therapeutic Chemical code classification, 2nd level: J06 |
| J07 | ATC | Vaccines | Number of prescriptions that match Anatomical Therapeutic Chemical code classification, 2nd level: J07 |
| L01 | ATC | Antineoplastic Agents | Number of prescriptions that match Anatomical Therapeutic Chemical code classification, 2nd level: L01 |
| L02 | ATC | Endocrine Therapy Antineoplastic And Immunomodulating Agents | Number of prescriptions that match Anatomical Therapeutic Chemical code classification, 2nd level: L02 |
| L03 | ATC | Immunostimulants | Number of prescriptions that match Anatomical Therapeutic Chemical code classification, 2nd level: L03 |
| L04 | ATC | Immunosuppressants | Number of prescriptions that match Anatomical Therapeutic Chemical code classification, 2nd level: L04 |
| M01 | ATC | Antiinflammatory And Antirheumatic Products | Number of prescriptions that match Anatomical Therapeutic Chemical code classification, 2nd level: M01 |

| | | | |
|---|---|---|---|
| M02 | ATC | Topical Products For Joint And Muscular Pain | Number of prescriptions that match Anatomical Therapeutic Chemical code classification, 2nd level: M02 |
| M03 | ATC | Muscle Relaxants | Number of prescriptions that match Anatomical Therapeutic Chemical code classification, 2nd level: M03 |
| M04 | ATC | Antigout Preparations | Number of prescriptions that match Anatomical Therapeutic Chemical code classification, 2nd level: M04 |
| M05 | ATC | Drugs For Treatment Of Bone Diseases | Number of prescriptions that match Anatomical Therapeutic Chemical code classification, 2nd level: M05 |
| M09 | ATC | Other Drugs For Disorders Of The Musculo-Skeletal System In Atc | Number of prescriptions that match Anatomical Therapeutic Chemical code classification, 2nd level: M09 |
| N01 | ATC | Anesthetics | Number of prescriptions that match Anatomical Therapeutic Chemical code classification, 2nd level: N01 |
| N02 | ATC | Analgesics | Number of prescriptions that match Anatomical Therapeutic Chemical code classification, 2nd level: N02 |
| N03 | ATC | Antiepileptics | Number of prescriptions that match Anatomical Therapeutic Chemical code classification, 2nd level: N03 |
| N04 | ATC | Anti-Parkinson Drugs | Number of prescriptions that match Anatomical Therapeutic Chemical code classification, 2nd level: N04 |
| N05 | ATC | Psycholeptics | Number of prescriptions that match Anatomical Therapeutic Chemical code classification, 2nd level: N05 |
| N06 | ATC | Psychoanaleptics | Number of prescriptions that match Anatomical Therapeutic Chemical code classification, 2nd level: N06 |
| N07 | ATC | Other Nervous System Drugs In Atc | Number of prescriptions that match Anatomical Therapeutic Chemical code classification, 2nd level: N07 |

| | | | |
|---|---|---|---|
| P01 | ATC | Antiprotozoals | Number of prescriptions that match Anatomical Therapeutic Chemical code classification, 2nd level: P01 |
| P02 | ATC | Anthelmintics | Number of prescriptions that match Anatomical Therapeutic Chemical code classification, 2nd level: P02 |
| P03 | ATC | Ectoparasiticides, Incl. Scabicides, Insecticides And Repellents | Number of prescriptions that match Anatomical Therapeutic Chemical code classification, 2nd level: P03 |
| R01 | ATC | Nasal Preparations | Number of prescriptions that match Anatomical Therapeutic Chemical code classification, 2nd level: R01 |
| R02 | ATC | Throat Preparations | Number of prescriptions that match Anatomical Therapeutic Chemical code classification, 2nd level: R02 |
| R03 | ATC | Drugs For Obstructive Airway Diseases | Number of prescriptions that match Anatomical Therapeutic Chemical code classification, 2nd level: R03 |
| R05 | ATC | Cough And Cold Preparations | Number of prescriptions that match Anatomical Therapeutic Chemical code classification, 2nd level: R05 |
| R06 | ATC | Antihistamines For Systemic Use | Number of prescriptions that match Anatomical Therapeutic Chemical code classification, 2nd level: R06 |
| R07 | ATC | Other Respiratory System Products In Atc | Number of prescriptions that match Anatomical Therapeutic Chemical code classification, 2nd level: R07 |
| S01 | ATC | Ophthalmologicals | Number of prescriptions that match Anatomical Therapeutic Chemical code classification, 2nd level: S01 |
| S02 | ATC | Otologicals | Number of prescriptions that match Anatomical Therapeutic Chemical code classification, 2nd level: S02 |
| S03 | ATC | Ophthalmological And Otological Preparations | Number of prescriptions that match Anatomical Therapeutic Chemical code classification, 2nd level: S03 |

| | | | |
|---|---|---|---|
| V03 | ATC | All Other Therapeutic Products | Number of prescriptions that match Anatomical Therapeutic Chemical code classification, 2nd level: V03 |
| V04 | ATC | Diagnostic Agents | Number of prescriptions that match Anatomical Therapeutic Chemical code classification, 2nd level: V04 |
| V06 | ATC | General Nutrients | Number of prescriptions that match Anatomical Therapeutic Chemical code classification, 2nd level: V06 |
| Anesthesia_1 | CPT | Anesthesia for Procedures on the Head | Number of times a Current Procedural Terminology code was documented in the group: Anesthesia for Procedures on the Head (CPT 100 - 222) |
| Anesthesia_10 | CPT | Anesthesia for Procedures on the Upper Leg (Except Knee) | Number of times a Current Procedural Terminology code was documented in the group: Anesthesia for Procedures on the Upper Leg (Except Knee) (CPT 1200 - 1274) |
| Anesthesia_11 | CPT | Anesthesia for Procedures on the Knee and Popliteal Area | Number of times a Current Procedural Terminology code was documented in the group: Anesthesia for Procedures on the Knee and Popliteal Area (CPT 1320 - 1444) |
| Anesthesia_12 | CPT | Anesthesia for Procedures on the Lower Leg (Below Knee) | Number of times a Current Procedural Terminology code was documented in the group: Anesthesia for Procedures on the Lower Leg (Below Knee) (CPT 1462 - 1522) |
| Anesthesia_13 | CPT | Anesthesia for Procedures on the Shoulder and Axilla | Number of times a Current Procedural Terminology code was documented in the group: Anesthesia for Procedures on the Shoulder and Axilla (CPT 1610 - 1680) |

| | | | |
|---|---|---|---|
| Anesthesia_14 | CPT | Anesthesia for Procedures on the Upper Arm and Elbow | Number of times a Current Procedural Terminology code was documented in the group: Anesthesia for Procedures on the Upper Arm and Elbow (CPT 1710 - 1782) |
| Anesthesia_15 | CPT | Anesthesia for Procedures on the Forearm, Wrist, and Hand | Number of times a Current Procedural Terminology code was documented in the group: Anesthesia for Procedures on the Forearm, Wrist, and Hand (CPT 1810 - 1860) |
| Anesthesia_16 | CPT | Anesthesia for Radiological Procedures | Number of times a Current Procedural Terminology code was documented in the group: Anesthesia for Radiological Procedures (CPT 1916 - 1936) |
| Anesthesia_18 | CPT | Anesthesia for Obstetric Procedures | Number of times a Current Procedural Terminology code was documented in the group: Anesthesia for Obstetric Procedures (CPT 1958 - 1969) |
| Anesthesia_19 | CPT | Anesthesia for Other Procedures | Number of times a Current Procedural Terminology code was documented in the group: Anesthesia for Other Procedures (CPT 1990 - 1999) |
| Anesthesia_2 | CPT | Anesthesia for Procedures on the Neck | Number of times a Current Procedural Terminology code was documented in the group: Anesthesia for Procedures on the Neck (CPT 300 - 352) |
| Anesthesia_3 | CPT | Anesthesia for Procedures on the Thorax (Chest Wall and Shoulder Girdle) | Number of times a Current Procedural Terminology code was documented in the group: Anesthesia for Procedures on the Thorax (Chest Wall and Shoulder Girdle) (CPT 400 - 474) |

| | | | |
|---|---|---|---|
| Anesthesia_4 | CPT | Anesthesia for Intrathoracic Procedures | Number of times a Current Procedural Terminology code was documented in the group: Anesthesia for Intrathoracic Procedures (CPT 500 - 580) |
| Anesthesia_5 | CPT | Anesthesia for Procedures on the Spine and Spinal Cord | Number of times a Current Procedural Terminology code was documented in the group: Anesthesia for Procedures on the Spine and Spinal Cord (CPT 600 - 670) |
| Anesthesia_6 | CPT | Anesthesia for Procedures on the Upper Abdomen | Number of times a Current Procedural Terminology code was documented in the group: Anesthesia for Procedures on the Upper Abdomen (CPT 700 - 797) |
| Anesthesia_7 | CPT | Anesthesia for Procedures on the Lower Abdomen | Number of times a Current Procedural Terminology code was documented in the group: Anesthesia for Procedures on the Lower Abdomen (CPT 800 - 882) |
| Anesthesia_8 | CPT | Anesthesia for Procedures on the Perineum | Number of times a Current Procedural Terminology code was documented in the group: Anesthesia for Procedures on the Perineum (CPT 902 - 952) |
| Anesthesia_9 | CPT | Anesthesia for Procedures on the Pelvis (Except Hip) | Number of times a Current Procedural Terminology code was documented in the group: Anesthesia for Procedures on the Pelvis (Except Hip) (CPT 1112 - 1173) |
| Evaluation_1 | CPT | Office or Other Outpatient Services | Number of times a Current Procedural Terminology code was documented in the group: Office or Other Outpatient Services (CPT 99201 - 99215) |
| Evaluation_11 | CPT | Prolonged Services | Number of times a Current Procedural Terminology code was documented in the group: Prolonged Services (CPT 99354 - 99416) |

| | | | |
|---|---|---|---|
| Evaluation_12 | CPT | Case Management Services | Number of times a Current Procedural Terminology code was documented in the group: Case Management Services (CPT 99366 - 99368) |
| Evaluation_13 | CPT | Care Plan Oversight Services | Number of times a Current Procedural Terminology code was documented in the group: Care Plan Oversight Services (CPT 99374 - 99380) |
| Evaluation_14 | CPT | Preventive Medicine Services | Number of times a Current Procedural Terminology code was documented in the group: Preventive Medicine Services (CPT 99381 - 99429) |
| Evaluation_15 | CPT | Non-Face-to-Face Evaluation and Management Services | Number of times a Current Procedural Terminology code was documented in the group: Non-Face-to-Face Evaluation and Management Services (CPT 99441 - 99458) |
| Evaluation_19 | CPT | Inpatient Neonatal Intensive Care Services and Pediatric and Neonatal Critical Care Services | Number of times a Current Procedural Terminology code was documented in the group: Inpatient Neonatal Intensive Care Services and Pediatric and Neonatal Critical Care Services (CPT 99466 - 99486) |
| Evaluation_2 | CPT | Hospital Observation Services | Number of times a Current Procedural Terminology code was documented in the group: Hospital Observation Services (CPT 99217 - 99226) |
| Evaluation_22 | CPT | Care Management Evaluation and Management Services | Number of times a Current Procedural Terminology code was documented in the group: Care Management Evaluation and Management Services (CPT 99487 - 99491) |

| | | | |
|---|---|---|---|
| Evaluation_24 | CPT | Transitional Care Evaluation and Management Services | Number of times a Current Procedural Terminology code was documented in the group: Transitional Care Evaluation and Management Services (CPT 99495 - 99496) |
| Evaluation_26 | CPT | Other Evaluation and Management Services | Number of times a Current Procedural Terminology code was documented in the group: Other Evaluation and Management Services (CPT 99499 - 99499) |
| Evaluation_3 | CPT | Hospital Inpatient Services | Number of times a Current Procedural Terminology code was documented in the group: Hospital Inpatient Services (CPT 99221 - 99239) |
| Evaluation_4 | CPT | Consultation Services | Number of times a Current Procedural Terminology code was documented in the group: Consultation Services (CPT 99241 - 99255) |
| Evaluation_5 | CPT | Emergency Department Services | Number of times a Current Procedural Terminology code was documented in the group: Emergency Department Services (CPT 99281 - 99288) |
| Evaluation_6 | CPT | Critical Care Services | Number of times a Current Procedural Terminology code was documented in the group: Critical Care Services (CPT 99291 - 99292) |
| Evaluation_7 | CPT | Nursing Facility Services | Number of times a Current Procedural Terminology code was documented in the group: Nursing Facility Services (CPT 99304 - 99318) |
| Evaluation_8 | CPT | Domiciliary, Rest Home (eg, Boarding Home), or Custodial Care Services | Number of times a Current Procedural Terminology code was documented in the group: Domiciliary, Rest Home (eg, Boarding Home), or Custodial Care Services (CPT 99324 - 99337) |

| | | | |
|---|---|---|---|
| Medical_10 | CPT | Cardiovascular Procedures | Number of times a Current Procedural Terminology code was documented in the group: Cardiovascular Procedures (CPT 92920 - 93799) |
| Medical_11 | CPT | Non-Invasive Vascular Diagnostic Studies | Number of times a Current Procedural Terminology code was documented in the group: Non-Invasive Vascular Diagnostic Studies (CPT 93880 - 93998) |
| Medical_12 | CPT | Pulmonary Procedures | Number of times a Current Procedural Terminology code was documented in the group: Pulmonary Procedures (CPT 94002 - 94799) |
| Medical_13 | CPT | Allergy and Clinical Immunology Procedures | Number of times a Current Procedural Terminology code was documented in the group: Allergy and Clinical Immunology Procedures (CPT 95004 - 95199) |
| Medical_14 | CPT | Endocrinology Services | Number of times a Current Procedural Terminology code was documented in the group: Endocrinology Services (CPT 95249 - 95251) |
| Medical_15 | CPT | Neurology and Neuromuscular Procedures | Number of times a Current Procedural Terminology code was documented in the group: Neurology and Neuromuscular Procedures (CPT 95700 - 96020) |
| Medical_16 | CPT | Medical Genetics and Genetic Counseling Services | Number of times a Current Procedural Terminology code was documented in the group: Medical Genetics and Genetic Counseling Services (CPT 96040 - 96040) |

| | | | |
|---|---|---|---|
| Medical_17 | CPT | Central Nervous System Assessments/Tests (eg, Neuro-Cognitive, Mental Status, Speech Testing) | Number of times a Current Procedural Terminology code was documented in the group: Central Nervous System Assessments/Tests (eg, Neuro-Cognitive, Mental Status, Speech Testing) (CPT 96105 - 96146) |
| Medical_19 | CPT | Hydration, Therapeutic, Prophylactic, Diagnostic Injections and Infusions, and Chemotherapy and Other Highly Complex Drug or Highly Complex Biologic Agent Administration | Number of times a Current Procedural Terminology code was documented in the group: Hydration, Therapeutic, Prophylactic, Diagnostic Injections and Infusions, and Chemotherapy and Other Highly Complex Drug or Highly Complex Biologic Agent Administration (CPT 96360 - 96549) |
| Medical_2 | CPT | Immunization Administration for Vaccines/Toxoids | Number of times a Current Procedural Terminology code was documented in the group: Immunization Administration for Vaccines/Toxoids (CPT 90460 - 90474) |
| Medical_20 | CPT | Photodynamic Therapy Procedures | Number of times a Current Procedural Terminology code was documented in the group: Photodynamic Therapy Procedures (CPT 96567 - 96574) |
| Medical_21 | CPT | Special Dermatological Procedures | Number of times a Current Procedural Terminology code was documented in the group: Special Dermatological Procedures (CPT 96900 - 96999) |
| Medical_23 | CPT | Physical Medicine and Rehabilitation Evaluations | Number of times a Current Procedural Terminology code was documented in the group: Physical Medicine and Rehabilitation Evaluations (CPT 97161 - 97799) |

| | | | |
|---|---|---|---|
| Medical_24 | CPT | Medical Nutrition Therapy Procedures | Number of times a Current Procedural Terminology code was documented in the group: Medical Nutrition Therapy Procedures (CPT 97802 - 97804) |
| Medical_25 | CPT | Acupuncture Procedures | Number of times a Current Procedural Terminology code was documented in the group: Acupuncture Procedures (CPT 97810 - 97814) |
| Medical_27 | CPT | Chiropractic Manipulative Treatment Procedures | Number of times a Current Procedural Terminology code was documented in the group: Chiropractic Manipulative Treatment Procedures (CPT 98940 - 98943) |
| Medical_28 | CPT | Education and Training for Patient Self-Management | Number of times a Current Procedural Terminology code was documented in the group: Education and Training for Patient Self-Management (CPT 98960 - 98962) |
| Medical_29 | CPT | Non-Face-to-Face Nonphysician Services | Number of times a Current Procedural Terminology code was documented in the group: Non-Face-to-Face Nonphysician Services (CPT 98966 - 98972) |
| Medical_3 | CPT | Vaccines, Toxoids | Number of times a Current Procedural Terminology code was documented in the group: Vaccines, Toxoids (CPT 90476 - 90756) |
| Medical_30 | CPT | Special Services, Procedures and Reports | Number of times a Current Procedural Terminology code was documented in the group: Special Services, Procedures and Reports (CPT 99000 - 99091) |

| | | | |
|---|---|---|---|
| Medical_31 | CPT | Qualifying Circumstances for Anesthesia | Number of times a Current Procedural Terminology code was documented in the group: Qualifying Circumstances for Anesthesia (CPT 99100 - 99140) |
| Medical_32 | CPT | Moderate (Conscious) Sedation | Number of times a Current Procedural Terminology code was documented in the group: Moderate (Conscious) Sedation (CPT 99151 - 99157) |
| Medical_33 | CPT | Other Medicine Services and Procedures | Number of times a Current Procedural Terminology code was documented in the group: Other Medicine Services and Procedures (CPT 99170 - 99199) |
| Medical_4 | CPT | Psychiatry Services and Procedures | Number of times a Current Procedural Terminology code was documented in the group: Psychiatry Services and Procedures (CPT 90785 - 90899) |
| Medical_5 | CPT | Biofeedback Services and Procedures | Number of times a Current Procedural Terminology code was documented in the group: Biofeedback Services and Procedures (CPT 90901 - 90913) |
| Medical_6 | CPT | Dialysis Services and Procedures | Number of times a Current Procedural Terminology code was documented in the group: Dialysis Services and Procedures (CPT 90935 - 90999) |
| Medical_7 | CPT | Gastroenterology Procedures | Number of times a Current Procedural Terminology code was documented in the group: Gastroenterology Procedures (CPT 91010 - 91299) |

| | | | |
|---|---|---|---|
| Medical_8 | CPT | Ophthalmology Services and Procedures | Number of times a Current Procedural Terminology code was documented in the group: Ophthalmology Services and Procedures (CPT 92002 - 92499) |
| Medical_9 | CPT | Special Otorhinolaryngologic Services and Procedures | Number of times a Current Procedural Terminology code was documented in the group: Special Otorhinolaryngologic Services and Procedures (CPT 92502 - 92700) |
| Pathology_10 | CPT | Drug Assay Procedures | Number of times a Current Procedural Terminology code was documented in the group: Drug Assay Procedures (CPT 80305 - 80377) |
| Pathology_12 | CPT | Clinical Pathology Consultations | Number of times a Current Procedural Terminology code was documented in the group: Clinical Pathology Consultations (CPT 80500 - 80502) |
| Pathology_13 | CPT | Urinalysis Procedures | Number of times a Current Procedural Terminology code was documented in the group: Urinalysis Procedures (CPT 81000 - 81099) |
| Pathology_14 | CPT | Molecular Pathology Procedures | Number of times a Current Procedural Terminology code was documented in the group: Molecular Pathology Procedures (CPT 81105 - 81408) |
| Pathology_15 | CPT | Genomic Sequencing Procedures and Other Molecular Multianalyte Assays | Number of times a Current Procedural Terminology code was documented in the group: Genomic Sequencing Procedures and Other Molecular Multianalyte Assays (CPT 81410 - 81479) |

| | | | |
|---|---|---|---|
| Pathology_17 | CPT | Chemistry Procedures | Number of times a Current Procedural Terminology code was documented in the group: Chemistry Procedures (CPT 82009 - 84999) |
| Pathology_18 | CPT | Hematology and Coagulation Procedures | Number of times a Current Procedural Terminology code was documented in the group: Hematology and Coagulation Procedures (CPT 85002 - 85999) |
| Pathology_19 | CPT | Immunology Procedures | Number of times a Current Procedural Terminology code was documented in the group: Immunology Procedures (CPT 86000 - 86849) |
| Pathology_20 | CPT | Transfusion Medicine Procedures | Number of times a Current Procedural Terminology code was documented in the group: Transfusion Medicine Procedures (CPT 86850 - 86999) |
| Pathology_21 | CPT | Microbiology Procedures | Number of times a Current Procedural Terminology code was documented in the group: Microbiology Procedures (CPT 87003 - 87999) |
| Pathology_23 | CPT | Cytopathology Procedures | Number of times a Current Procedural Terminology code was documented in the group: Cytopathology Procedures (CPT 88104 - 88199) |
| Pathology_24 | CPT | Cytogenetic Studies | Number of times a Current Procedural Terminology code was documented in the group: Cytogenetic Studies (CPT 88230 - 88299) |
| Pathology_25 | CPT | Surgical Pathology Procedures | Number of times a Current Procedural Terminology code was documented in the group: Surgical Pathology Procedures (CPT 88300 - 88399) |

| | | | |
|---|---|---|---|
| Pathology_27 | CPT | Other Pathology and Laboratory Procedures | Number of times a Current Procedural Terminology code was documented in the group: Other Pathology and Laboratory Procedures (CPT 89049 - 89240) |
| Pathology_28 | CPT | Reproductive Medicine Procedures | Number of times a Current Procedural Terminology code was documented in the group: Reproductive Medicine Procedures (CPT 89250 - 89398) |
| Pathology_8 | CPT | Organ or Disease Oriented Panels | Number of times a Current Procedural Terminology code was documented in the group: Organ or Disease Oriented Panels (CPT 80047 - 80081) |
| Pathology_9 | CPT | Therapeutic Drug Assays | Number of times a Current Procedural Terminology code was documented in the group: Therapeutic Drug Assays (CPT 80145 - 80377) |
| Radiology_1 | CPT | Diagnostic Radiology (Diagnostic Imaging) Procedures | Number of times a Current Procedural Terminology code was documented in the group: Diagnostic Radiology (Diagnostic Imaging) Procedures (CPT 70010 - 76499) |
| Radiology_2 | CPT | Diagnostic Ultrasound Procedures | Number of times a Current Procedural Terminology code was documented in the group: Diagnostic Ultrasound Procedures (CPT 76506 - 76999) |
| Radiology_3 | CPT | Radiologic Guidance | Number of times a Current Procedural Terminology code was documented in the group: Radiologic Guidance (CPT 77001 - 77022) |
| Radiology_4 | CPT | Breast, Mammography | Number of times a Current Procedural Terminology code was documented in the group: Breast, Mammography (CPT 77046 - 77067) |

| | | | |
|---|---|---|---|
| Radiology_5 | CPT | Bone/Joint Studies | Number of times a Current Procedural Terminology code was documented in the group: Bone/Joint Studies (CPT 77071 - 77086) |
| Radiology_6 | CPT | Radiation Oncology Treatment | Number of times a Current Procedural Terminology code was documented in the group: Radiation Oncology Treatment (CPT 77261 - 77799) |
| Radiology_7 | CPT | Nuclear Medicine Procedures | Number of times a Current Procedural Terminology code was documented in the group: Nuclear Medicine Procedures (CPT 78012 - 79999) |
| Surgery_1 | CPT | General Surgical Procedures | Number of times a Current Procedural Terminology code was documented in the group: General Surgical Procedures (CPT 10004 - 10021) |
| Surgery_10 | CPT | Surgical Procedures on the Male Genital System | Number of times a Current Procedural Terminology code was documented in the group: Surgical Procedures on the Male Genital System (CPT 54000 - 55899) |
| Surgery_11 | CPT | Reproductive System Procedures | Number of times a Current Procedural Terminology code was documented in the group: Reproductive System Procedures (CPT 55920 - 55920) |
| Surgery_13 | CPT | Surgical Procedures on the Female Genital System | Number of times a Current Procedural Terminology code was documented in the group: Surgical Procedures on the Female Genital System (CPT 56405 - 58999) |

| | | | |
|---|---|---|---|
| Surgery_14 | CPT | Surgical Procedures for Maternity Care and Delivery | Number of times a Current Procedural Terminology code was documented in the group: Surgical Procedures for Maternity Care and Delivery (CPT 59000 - 59899) |
| Surgery_15 | CPT | Surgical Procedures on the Endocrine System | Number of times a Current Procedural Terminology code was documented in the group: Surgical Procedures on the Endocrine System (CPT 60000 - 60699) |
| Surgery_16 | CPT | Surgical Procedures on the Nervous System | Number of times a Current Procedural Terminology code was documented in the group: Surgical Procedures on the Nervous System (CPT 61000 - 64999) |
| Surgery_17 | CPT | Surgical Procedures on the Eye and Ocular Adnexa | Number of times a Current Procedural Terminology code was documented in the group: Surgical Procedures on the Eye and Ocular Adnexa (CPT 65091 - 68899) |
| Surgery_18 | CPT | Surgical Procedures on the Auditory System | Number of times a Current Procedural Terminology code was documented in the group: Surgical Procedures on the Auditory System (CPT 69000 - 69979) |
| Surgery_19 | CPT | Operating Microscope Procedures | Number of times a Current Procedural Terminology code was documented in the group: Operating Microscope Procedures (CPT 69990 - 69990) |
| Surgery_2 | CPT | Surgical Procedures on the Integumentary System | Number of times a Current Procedural Terminology code was documented in the group: Surgical Procedures on the Integumentary System (CPT 10030 - 19499) |

| | | | |
|---|---|---|---|
| Surgery_3 | CPT | Surgical Procedures on the Musculoskeletal System | Number of times a Current Procedural Terminology code was documented in the group: Surgical Procedures on the Musculoskeletal System (CPT 20100 - 29999) |
| Surgery_4 | CPT | Surgical Procedures on the Respiratory System | Number of times a Current Procedural Terminology code was documented in the group: Surgical Procedures on the Respiratory System (CPT 30000 - 32999) |
| Surgery_5 | CPT | Surgical Procedures on the Cardiovascular System | Number of times a Current Procedural Terminology code was documented in the group: Surgical Procedures on the Cardiovascular System (CPT 33016 - 37799) |
| Surgery_6 | CPT | Surgical Procedures on the Hemic and Lymphatic Systems | Number of times a Current Procedural Terminology code was documented in the group: Surgical Procedures on the Hemic and Lymphatic Systems (CPT 38100 - 38999) |
| Surgery_7 | CPT | Surgical Procedures on the Mediastinum and Diaphragm | Number of times a Current Procedural Terminology code was documented in the group: Surgical Procedures on the Mediastinum and Diaphragm (CPT 39000 - 39599) |
| Surgery_8 | CPT | Surgical Procedures on the Digestive System | Number of times a Current Procedural Terminology code was documented in the group: Surgical Procedures on the Digestive System (CPT 40490 - 49999) |
| Surgery_9 | CPT | Surgical Procedures on the Urinary System | Number of times a Current Procedural Terminology code was documented in the group: Surgical Procedures on the Urinary System (CPT 50010 - 53899) |
| age_at_scan | Demographics | Age | Age at time of abdominal scan in years |

| | | | |
|---|---|---|---|
| gender | Demographics | Male | Whether the patient is male |
| Chapter_I_1 | ICD10 | Intestinal infectious diseases | Number of times an International Classification of Diseases, 10th edition was documented in the year prior to imaging: Intestinal infectious diseases (ICD10 Codes A00 - A09) |
| Chapter_I_11 | ICD10 | Viral infections characterized by skin and mucous membrane lesions | Number of times an International Classification of Diseases, 10th edition was documented in the year prior to imaging: Viral infections characterized by skin and mucous membrane lesions (ICD10 Codes B00 - B09) |
| Chapter_I_12 | ICD10 | Viral hepatitis | Number of times an International Classification of Diseases, 10th edition was documented in the year prior to imaging: Viral hepatitis (ICD10 Codes B15 - B19) |
| Chapter_I_13 | ICD10 | Human immunodeficiency virus [HIV] disease | Number of times an International Classification of Diseases, 10th edition was documented in the year prior to imaging: Human immunodeficiency virus [HIV] disease (ICD10 Codes B20 - B24) |
| Chapter_I_14 | ICD10 | Other viral diseases | Number of times an International Classification of Diseases, 10th edition was documented in the year prior to imaging: Other viral diseases (ICD10 Codes B25 - B34) |
| Chapter_I_15 | ICD10 | Mycoses | Number of times an International Classification of Diseases, 10th edition was documented in the year prior to imaging: Mycoses (ICD10 Codes B35 - B49) |

| | | | |
|---|---|---|---|
| Chapter_I_16 | ICD10 | Protozoal diseases | Number of times an International Classification of Diseases, 10th edition was documented in the year prior to imaging: Protozoal diseases (ICD10 Codes B50 - B64) |
| Chapter_I_17 | ICD10 | Helminthiases | Number of times an International Classification of Diseases, 10th edition was documented in the year prior to imaging: Helminthiases (ICD10 Codes B65 - B83) |
| Chapter_I_18 | ICD10 | Pediculosis, acariasis and other infestations | Number of times an International Classification of Diseases, 10th edition was documented in the year prior to imaging: Pediculosis, acariasis and other infestations (ICD10 Codes B85 - B89) |
| Chapter_I_19 | ICD10 | Sequelae of infectious and parasitic diseases | Number of times an International Classification of Diseases, 10th edition was documented in the year prior to imaging: Sequelae of infectious and parasitic diseases (ICD10 Codes B90 - B94) |
| Chapter_I_2 | ICD10 | Tuberculosis | Number of times an International Classification of Diseases, 10th edition was documented in the year prior to imaging: Tuberculosis (ICD10 Codes A15 - A19) |
| Chapter_I_20 | ICD10 | Bacterial, viral and other infectious agents | Number of times an International Classification of Diseases, 10th edition was documented in the year prior to imaging: Bacterial, viral and other infectious agents (ICD10 Codes B95 - B98) |

| | | | |
|---|---|---|---|
| Chapter_I_21 | ICD10 | Other infectious diseases | Number of times an International Classification of Diseases, 10th edition was documented in the year prior to imaging: Other infectious diseases (ICD10 Codes B99 - B99) |
| Chapter_I_3 | ICD10 | Certain zoonotic bacterial diseases | Number of times an International Classification of Diseases, 10th edition was documented in the year prior to imaging: Certain zoonotic bacterial diseases (ICD10 Codes A20 - A28) |
| Chapter_I_4 | ICD10 | Other bacterial diseases | Number of times an International Classification of Diseases, 10th edition was documented in the year prior to imaging: Other bacterial diseases (ICD10 Codes A30 - A49) |
| Chapter_I_5 | ICD10 | Infections with a predominantly sexual mode of transmission | Number of times an International Classification of Diseases, 10th edition was documented in the year prior to imaging: Infections with a predominantly sexual mode of transmission (ICD10 Codes A50 - A64) |
| Chapter_I_6 | ICD10 | Other spirochaetal diseases | Number of times an International Classification of Diseases, 10th edition was documented in the year prior to imaging: Other spirochaetal diseases (ICD10 Codes A65 - A69) |
| Chapter_I_7 | ICD10 | Other diseases caused by chlamydiae | Number of times an International Classification of Diseases, 10th edition was documented in the year prior to imaging: Other diseases caused by chlamydiae (ICD10 Codes A70 - A74) |

| | | | |
|---|---|---|---|
| Chapter_I_8 | ICD10 | Rickettsioses | Number of times an International Classification of Diseases, 10th edition was documented in the year prior to imaging: Rickettsioses (ICD10 Codes A75 - A79) |
| Chapter_I_9 | ICD10 | Viral infections of the central nervous system | Number of times an International Classification of Diseases, 10th edition was documented in the year prior to imaging: Viral infections of the central nervous system (ICD10 Codes A80 - A89) |
| Chapter_II_1 | ICD10 | Malignant neoplasms | Number of times an International Classification of Diseases, 10th edition was documented in the year prior to imaging: Malignant neoplasms (ICD10 Codes C00 - C97) |
| Chapter_II_2 | ICD10 | In situ neoplasms | Number of times an International Classification of Diseases, 10th edition was documented in the year prior to imaging: In situ neoplasms (ICD10 Codes D00 - D09) |
| Chapter_II_3 | ICD10 | Benign neoplasms | Number of times an International Classification of Diseases, 10th edition was documented in the year prior to imaging: Benign neoplasms (ICD10 Codes D10 - D36) |
| Chapter_II_4 | ICD10 | Neoplasms of uncertain or unknown behaviour | Number of times an International Classification of Diseases, 10th edition was documented in the year prior to imaging: Neoplasms of uncertain or unknown behaviour (ICD10 Codes D37 - D48) |
| Chapter_III_1 | ICD10 | Nutritional anaemias | Number of times an International Classification of Diseases, 10th edition was documented in the year prior to imaging: Nutritional anaemias (ICD10 Codes D50 - D53) |

| | | | |
|---|---|---|---|
| Chapter_III_2 | ICD10 | Haemolytic anaemias | Number of times an International Classification of Diseases, 10th edition was documented in the year prior to imaging: Haemolytic anaemias (ICD10 Codes D55 - D59) |
| Chapter_III_3 | ICD10 | Aplastic and other anaemias | Number of times an International Classification of Diseases, 10th edition was documented in the year prior to imaging: Aplastic and other anaemias (ICD10 Codes D60 - D64) |
| Chapter_III_4 | ICD10 | Coagulation defects, purpura and other haemorrhagic conditions | Number of times an International Classification of Diseases, 10th edition was documented in the year prior to imaging: Coagulation defects, purpura and other haemorrhagic conditions (ICD10 Codes D65 - D69) |
| Chapter_III_5 | ICD10 | Other diseases of blood and blood-forming organs | Number of times an International Classification of Diseases, 10th edition was documented in the year prior to imaging: Other diseases of blood and blood-forming organs (ICD10 Codes D70 - D77) |
| Chapter_III_6 | ICD10 | Certain disorders involving the immune mechanism | Number of times an International Classification of Diseases, 10th edition was documented in the year prior to imaging: Certain disorders involving the immune mechanism (ICD10 Codes D80 - D89) |
| Chapter_IV_1 | ICD10 | Disorders of thyroid gland | Number of times an International Classification of Diseases, 10th edition was documented in the year prior to imaging: Disorders of thyroid gland (ICD10 Codes E00 - E07) |

| | | | |
|---|---|---|---|
| Chapter_IV_2 | ICD10 | Diabetes mellitus | Number of times an International Classification of Diseases, 10th edition was documented in the year prior to imaging: Diabetes mellitus (ICD10 Codes E10 - E14) |
| Chapter_IV_3 | ICD10 | Other disorders of glucose regulation and pancreatic internal secretion | Number of times an International Classification of Diseases, 10th edition was documented in the year prior to imaging: Other disorders of glucose regulation and pancreatic internal secretion (ICD10 Codes E15 - E16) |
| Chapter_IV_4 | ICD10 | Disorders of other endocrine glands | Number of times an International Classification of Diseases, 10th edition was documented in the year prior to imaging: Disorders of other endocrine glands (ICD10 Codes E20 - E35) |
| Chapter_IV_5 | ICD10 | Malnutrition | Number of times an International Classification of Diseases, 10th edition was documented in the year prior to imaging: Malnutrition (ICD10 Codes E40 - E46) |
| Chapter_IV_6 | ICD10 | Other nutritional deficiencies | Number of times an International Classification of Diseases, 10th edition was documented in the year prior to imaging: Other nutritional deficiencies (ICD10 Codes E50 - E64) |
| Chapter_IV_7 | ICD10 | Obesity and other hyperalimentation | Number of times an International Classification of Diseases, 10th edition was documented in the year prior to imaging: Obesity and other hyperalimentation (ICD10 Codes E65 - E68) |
| Chapter_IV_8 | ICD10 | Metabolic disorders | Number of times an International Classification of Diseases, 10th edition was documented in the year prior to imaging: Metabolic disorders (ICD10 Codes E70 - E90) |

| | | | |
|---|---|---|---|
| Chapter_IX_1 | ICD10 | Acute rheumatic fever | Number of times an International Classification of Diseases, 10th edition was documented in the year prior to imaging: Acute rheumatic fever (ICD10 Codes I00 - I02) |
| Chapter_IX_10 | ICD10 | Other and unspecified disorders of the circulatory system | Number of times an International Classification of Diseases, 10th edition was documented in the year prior to imaging: Other and unspecified disorders of the circulatory system (ICD10 Codes I95 - I99) |
| Chapter_IX_2 | ICD10 | Chronic rheumatic heart diseases | Number of times an International Classification of Diseases, 10th edition was documented in the year prior to imaging: Chronic rheumatic heart diseases (ICD10 Codes I05 - I09) |
| Chapter_IX_3 | ICD10 | Hypertensive diseases | Number of times an International Classification of Diseases, 10th edition was documented in the year prior to imaging: Hypertensive diseases (ICD10 Codes I10 - I15) |
| Chapter_IX_4 | ICD10 | Ischaemic heart diseases | Number of times an International Classification of Diseases, 10th edition was documented in the year prior to imaging: Ischaemic heart diseases (ICD10 Codes I20 - I25) |
| Chapter_IX_5 | ICD10 | Pulmonary heart disease and diseases of pulmonary circulation | Number of times an International Classification of Diseases, 10th edition was documented in the year prior to imaging: Pulmonary heart disease and diseases of pulmonary circulation (ICD10 Codes I26 - I28) |

| | | | |
|---|---|---|---|
| Chapter_IX_6 | ICD10 | Other forms of heart disease | Number of times an International Classification of Diseases, 10th edition was documented in the year prior to imaging: Other forms of heart disease (ICD10 Codes I30 - I52) |
| Chapter_IX_7 | ICD10 | Cerebrovascular diseases | Number of times an International Classification of Diseases, 10th edition was documented in the year prior to imaging: Cerebrovascular diseases (ICD10 Codes I60 - I69) |
| Chapter_IX_8 | ICD10 | Diseases of arteries, arterioles and capillaries | Number of times an International Classification of Diseases, 10th edition was documented in the year prior to imaging: Diseases of arteries, arterioles and capillaries (ICD10 Codes I70 - I79) |
| Chapter_IX_9 | ICD10 | Diseases of veins, lymphatic vessels and lymph nodes, not elsewhere classified | Number of times an International Classification of Diseases, 10th edition was documented in the year prior to imaging: Diseases of veins, lymphatic vessels and lymph nodes, not elsewhere classified (ICD10 Codes I80 - I89) |
| Chapter_V_1 | ICD10 | Organic, including symptomatic, mental disorders | Number of times an International Classification of Diseases, 10th edition was documented in the year prior to imaging: Organic, including symptomatic, mental disorders (ICD10 Codes F00 - F09) |
| Chapter_V_10 | ICD10 | Behavioural and emotional disorders with onset usually occurring in childhood and adolescence | Number of times an International Classification of Diseases, 10th edition was documented in the year prior to imaging: Behavioural and emotional disorders with onset usually occurring in childhood and adolescence (ICD10 Codes F90 - F98) |

| | | | |
|---|---|---|---|
| Chapter_V_11 | ICD10 | Unspecified mental disorder | Number of times an International Classification of Diseases, 10th edition was documented in the year prior to imaging: Unspecified mental disorder (ICD10 Codes F99 - F99) |
| Chapter_V_2 | ICD10 | Mental and behavioural disorders due to psychoactive substance use | Number of times an International Classification of Diseases, 10th edition was documented in the year prior to imaging: Mental and behavioural disorders due to psychoactive substance use (ICD10 Codes F10 - F19) |
| Chapter_V_3 | ICD10 | Schizophrenia, schizotypal and delusional disorders | Number of times an International Classification of Diseases, 10th edition was documented in the year prior to imaging: Schizophrenia, schizotypal and delusional disorders (ICD10 Codes F20 - F29) |
| Chapter_V_4 | ICD10 | Mood [affective] disorders | Number of times an International Classification of Diseases, 10th edition was documented in the year prior to imaging: Mood [affective] disorders (ICD10 Codes F30 - F39) |
| Chapter_V_5 | ICD10 | Neurotic, stress-related and somatoform disorders | Number of times an International Classification of Diseases, 10th edition was documented in the year prior to imaging: Neurotic, stress-related and somatoform disorders (ICD10 Codes F40 - F48) |
| Chapter_V_6 | ICD10 | Behavioural syndromes associated with physiological disturbances and physical factors | Number of times an International Classification of Diseases, 10th edition was documented in the year prior to imaging: Behavioural syndromes associated with physiological disturbances and physical factors (ICD10 Codes F50 - F59) |

| | | | |
|---|---|---|---|
| Chapter_V_7 | ICD10 | Disorders of adult personality and behaviour | Number of times an International Classification of Diseases, 10th edition was documented in the year prior to imaging: Disorders of adult personality and behaviour (ICD10 Codes F60 - F69) |
| Chapter_V_8 | ICD10 | Mental retardation | Number of times an International Classification of Diseases, 10th edition was documented in the year prior to imaging: Mental retardation (ICD10 Codes F70 - F79) |
| Chapter_V_9 | ICD10 | Disorders of psychological development | Number of times an International Classification of Diseases, 10th edition was documented in the year prior to imaging: Disorders of psychological development (ICD10 Codes F80 - F89) |
| Chapter_VI_1 | ICD10 | Inflammatory diseases of the central nervous system | Number of times an International Classification of Diseases, 10th edition was documented in the year prior to imaging: Inflammatory diseases of the central nervous system (ICD10 Codes G00 - G09) |
| Chapter_VI_10 | ICD10 | Cerebral palsy and other paralytic syndromes | Number of times an International Classification of Diseases, 10th edition was documented in the year prior to imaging: Cerebral palsy and other paralytic syndromes (ICD10 Codes G80 - G83) |
| Chapter_VI_11 | ICD10 | Other disorders of the nervous system | Number of times an International Classification of Diseases, 10th edition was documented in the year prior to imaging: Other disorders of the nervous system (ICD10 Codes G90 - G99) |

| | | | |
|---|---|---|---|
| Chapter_VI_2 | ICD10 | Systemic atrophies primarily affecting the central nervous system | Number of times an International Classification of Diseases, 10th edition was documented in the year prior to imaging: Systemic atrophies primarily affecting the central nervous system (ICD10 Codes G10 - G14) |
| Chapter_VI_3 | ICD10 | Extrapyramidal and movement disorders | Number of times an International Classification of Diseases, 10th edition was documented in the year prior to imaging: Extrapyramidal and movement disorders (ICD10 Codes G20 - G26) |
| Chapter_VI_4 | ICD10 | Other degenerative diseases of the nervous system | Number of times an International Classification of Diseases, 10th edition was documented in the year prior to imaging: Other degenerative diseases of the nervous system (ICD10 Codes G30 - G32) |
| Chapter_VI_5 | ICD10 | Demyelinating diseases of the central nervous system | Number of times an International Classification of Diseases, 10th edition was documented in the year prior to imaging: Demyelinating diseases of the central nervous system (ICD10 Codes G35 - G37) |
| Chapter_VI_6 | ICD10 | Episodic and paroxysmal disorders | Number of times an International Classification of Diseases, 10th edition was documented in the year prior to imaging: Episodic and paroxysmal disorders (ICD10 Codes G40 - G47) |
| Chapter_VI_7 | ICD10 | Nerve, nerve root and plexus disorders | Number of times an International Classification of Diseases, 10th edition was documented in the year prior to imaging: Nerve, nerve root and plexus disorders (ICD10 Codes G50 - G59) |

| | | | |
|---|---|---|---|
| Chapter_VI_8 | ICD10 | Polyneuropathies and other disorders of the peripheral nervous system | Number of times an International Classification of Diseases, 10th edition was documented in the year prior to imaging: Polyneuropathies and other disorders of the peripheral nervous system (ICD10 Codes G60 - G64) |
| Chapter_VI_9 | ICD10 | Diseases of myoneural junction and muscle | Number of times an International Classification of Diseases, 10th edition was documented in the year prior to imaging: Diseases of myoneural junction and muscle (ICD10 Codes G70 - G73) |
| Chapter_VII_1 | ICD10 | Disorders of eyelid, lacrimal system and orbit | Number of times an International Classification of Diseases, 10th edition was documented in the year prior to imaging: Disorders of eyelid, lacrimal system and orbit (ICD10 Codes H00 - H06) |
| Chapter_VII_10 | ICD10 | Visual disturbances and blindness | Number of times an International Classification of Diseases, 10th edition was documented in the year prior to imaging: Visual disturbances and blindness (ICD10 Codes H53 - H54) |
| Chapter_VII_11 | ICD10 | Other disorders of eye and adnexa | Number of times an International Classification of Diseases, 10th edition was documented in the year prior to imaging: Other disorders of eye and adnexa (ICD10 Codes H55 - H59) |
| Chapter_VII_2 | ICD10 | Disorders of conjunctiva | Number of times an International Classification of Diseases, 10th edition was documented in the year prior to imaging: Disorders of conjunctiva (ICD10 Codes H10 - H13) |

| | | | |
|---|---|---|---|
| Chapter_VII_3 | ICD10 | Disorders of sclera, cornea, iris and ciliary body | Number of times an International Classification of Diseases, 10th edition was documented in the year prior to imaging: Disorders of sclera, cornea, iris and ciliary body (ICD10 Codes H15 - H22) |
| Chapter_VII_4 | ICD10 | Disorders of lens | Number of times an International Classification of Diseases, 10th edition was documented in the year prior to imaging: Disorders of lens (ICD10 Codes H25 - H28) |
| Chapter_VII_5 | ICD10 | Disorders of choroid and retina | Number of times an International Classification of Diseases, 10th edition was documented in the year prior to imaging: Disorders of choroid and retina (ICD10 Codes H30 - H36) |
| Chapter_VII_6 | ICD10 | Glaucoma | Number of times an International Classification of Diseases, 10th edition was documented in the year prior to imaging: Glaucoma (ICD10 Codes H40 - H42) |
| Chapter_VII_7 | ICD10 | Disorders of vitreous body and globe | Number of times an International Classification of Diseases, 10th edition was documented in the year prior to imaging: Disorders of vitreous body and globe (ICD10 Codes H43 - H45) |
| Chapter_VII_8 | ICD10 | Disorders of optic nerve and visual pathways | Number of times an International Classification of Diseases, 10th edition was documented in the year prior to imaging: Disorders of optic nerve and visual pathways (ICD10 Codes H46 - H48) |

| | | | |
|---|---|---|---|
| Chapter_VII_9 | ICD10 | Disorders of ocular muscles, binocular movement, accommodation and refraction | Number of times an International Classification of Diseases, 10th edition was documented in the year prior to imaging: Disorders of ocular muscles, binocular movement, accommodation and refraction (ICD10 Codes H49 - H52) |
| Chapter_VIII_1 | ICD10 | Diseases of external ear | Number of times an International Classification of Diseases, 10th edition was documented in the year prior to imaging: Diseases of external ear (ICD10 Codes H60 - H62) |
| Chapter_VIII_2 | ICD10 | Diseases of middle ear and mastoid | Number of times an International Classification of Diseases, 10th edition was documented in the year prior to imaging: Diseases of middle ear and mastoid (ICD10 Codes H65 - H75) |
| Chapter_VIII_3 | ICD10 | Diseases of inner ear | Number of times an International Classification of Diseases, 10th edition was documented in the year prior to imaging: Diseases of inner ear (ICD10 Codes H80 - H83) |
| Chapter_VIII_4 | ICD10 | Other disorders of ear | Number of times an International Classification of Diseases, 10th edition was documented in the year prior to imaging: Other disorders of ear (ICD10 Codes H90 - H95) |
| Chapter_X_1 | ICD10 | Acute upper respiratory infections | Number of times an International Classification of Diseases, 10th edition was documented in the year prior to imaging: Acute upper respiratory infections (ICD10 Codes J00 - J06) |

| | | | |
|---|---|---|---|
| Chapter_X_10 | ICD10 | Other diseases of the respiratory system | Number of times an International Classification of Diseases, 10th edition was documented in the year prior to imaging: Other diseases of the respiratory system (ICD10 Codes J95 - J99) |
| Chapter_X_2 | ICD10 | Influenza and pneumonia | Number of times an International Classification of Diseases, 10th edition was documented in the year prior to imaging: Influenza and pneumonia (ICD10 Codes J09 - J18) |
| Chapter_X_3 | ICD10 | Other acute lower respiratory infections | Number of times an International Classification of Diseases, 10th edition was documented in the year prior to imaging: Other acute lower respiratory infections (ICD10 Codes J20 - J22) |
| Chapter_X_4 | ICD10 | Other diseases of upper respiratory tract | Number of times an International Classification of Diseases, 10th edition was documented in the year prior to imaging: Other diseases of upper respiratory tract (ICD10 Codes J30 - J39) |
| Chapter_X_5 | ICD10 | Chronic lower respiratory diseases | Number of times an International Classification of Diseases, 10th edition was documented in the year prior to imaging: Chronic lower respiratory diseases (ICD10 Codes J40 - J47) |
| Chapter_X_6 | ICD10 | Lung diseases due to external agents | Number of times an International Classification of Diseases, 10th edition was documented in the year prior to imaging: Lung diseases due to external agents (ICD10 Codes J60 - J70) |

| | | | |
|---|---|---|---|
| Chapter_X_7 | ICD10 | Other respiratory diseases principally affecting the interstitium | Number of times an International Classification of Diseases, 10th edition was documented in the year prior to imaging: Other respiratory diseases principally affecting the interstitium (ICD10 Codes J80 - J84) |
| Chapter_X_8 | ICD10 | Suppurative and necrotic conditions of lower respiratory tract | Number of times an International Classification of Diseases, 10th edition was documented in the year prior to imaging: Suppurative and necrotic conditions of lower respiratory tract (ICD10 Codes J85 - J86) |
| Chapter_X_9 | ICD10 | Other diseases of pleura | Number of times an International Classification of Diseases, 10th edition was documented in the year prior to imaging: Other diseases of pleura (ICD10 Codes J90 - J94) |
| Chapter_XI_1 | ICD10 | Diseases of oral cavity, salivary glands and jaws | Number of times an International Classification of Diseases, 10th edition was documented in the year prior to imaging: Diseases of oral cavity, salivary glands and jaws (ICD10 Codes K00 - K14) |
| Chapter_XI_10 | ICD10 | Other diseases of the digestive system | Number of times an International Classification of Diseases, 10th edition was documented in the year prior to imaging: Other diseases of the digestive system (ICD10 Codes K90 - K93) |
| Chapter_XI_2 | ICD10 | Bullous disorders | Number of times an International Classification of Diseases, 10th edition was documented in the year prior to imaging: Bullous disorders (ICD10 Codes L10 - L14) |

| | | | |
|---|---|---|---|
| Chapter_XI_3 | ICD10 | Dermatitis and eczema | Number of times an International Classification of Diseases, 10th edition was documented in the year prior to imaging: Dermatitis and eczema (ICD10 Codes L20 - L30) |
| Chapter_XI_4 | ICD10 | Papulosquamous disorders | Number of times an International Classification of Diseases, 10th edition was documented in the year prior to imaging: Papulosquamous disorders (ICD10 Codes L40 - L45) |
| Chapter_XI_5 | ICD10 | Noninfective enteritis and colitis | Number of times an International Classification of Diseases, 10th edition was documented in the year prior to imaging: Noninfective enteritis and colitis (ICD10 Codes K50 - K52) |
| Chapter_XI_6 | ICD10 | Other diseases of intestines | Number of times an International Classification of Diseases, 10th edition was documented in the year prior to imaging: Other diseases of intestines (ICD10 Codes K55 - K64) |
| Chapter_XI_7 | ICD10 | Diseases of peritoneum | Number of times an International Classification of Diseases, 10th edition was documented in the year prior to imaging: Diseases of peritoneum (ICD10 Codes K65 - K67) |
| Chapter_XI_8 | ICD10 | Diseases of liver | Number of times an International Classification of Diseases, 10th edition was documented in the year prior to imaging: Diseases of liver (ICD10 Codes K70 - K77) |

| | | | |
|---|---|---|---|
| Chapter_XI_9 | ICD10 | Disorders of gallbladder, biliary tract and pancreas | Number of times an International Classification of Diseases, 10th edition was documented in the year prior to imaging: Disorders of gallbladder, biliary tract and pancreas (ICD10 Codes K80 - K87) |
| Chapter_XII_1 | ICD10 | Infections of the skin and subcutaneous tissue | Number of times an International Classification of Diseases, 10th edition was documented in the year prior to imaging: Infections of the skin and subcutaneous tissue (ICD10 Codes L00 - L08) |
| Chapter_XII_2 | ICD10 | Bullous disorders | Number of times an International Classification of Diseases, 10th edition was documented in the year prior to imaging: Bullous disorders (ICD10 Codes L10 - L14) |
| Chapter_XII_3 | ICD10 | Dermatitis and eczema | Number of times an International Classification of Diseases, 10th edition was documented in the year prior to imaging: Dermatitis and eczema (ICD10 Codes L20 - L30) |
| Chapter_XII_4 | ICD10 | Papulosquamous disorders | Number of times an International Classification of Diseases, 10th edition was documented in the year prior to imaging: Papulosquamous disorders (ICD10 Codes L40 - L45) |
| Chapter_XII_5 | ICD10 | Urticaria and erythema | Number of times an International Classification of Diseases, 10th edition was documented in the year prior to imaging: Urticaria and erythema (ICD10 Codes L50 - L54) |

| | | | |
|---|---|---|---|
| Chapter_XII_6 | ICD10 | Radiation-related disorders of the skin and subcutaneous tissue | Number of times an International Classification of Diseases, 10th edition was documented in the year prior to imaging: Radiation-related disorders of the skin and subcutaneous tissue (ICD10 Codes L55 - L59) |
| Chapter_XII_7 | ICD10 | Disorders of skin appendages | Number of times an International Classification of Diseases, 10th edition was documented in the year prior to imaging: Disorders of skin appendages (ICD10 Codes L60 - L75) |
| Chapter_XII_8 | ICD10 | Other disorders of the skin and subcutaneous tissue | Number of times an International Classification of Diseases, 10th edition was documented in the year prior to imaging: Other disorders of the skin and subcutaneous tissue (ICD10 Codes L80 - L99) |
| Chapter_XIII_1 | ICD10 | Arthropathies | Number of times an International Classification of Diseases, 10th edition was documented in the year prior to imaging: Arthropathies (ICD10 Codes M00 - M25) |
| Chapter_XIII_2 | ICD10 | Systemic connective tissue disorders | Number of times an International Classification of Diseases, 10th edition was documented in the year prior to imaging: Systemic connective tissue disorders (ICD10 Codes M30 - M36) |
| Chapter_XIII_3 | ICD10 | Dorsopathies | Number of times an International Classification of Diseases, 10th edition was documented in the year prior to imaging: Dorsopathies (ICD10 Codes M40 - M54) |

| | | | |
|---|---|---|---|
| Chapter_XIII_4 | ICD10 | Soft tissue disorders | Number of times an International Classification of Diseases, 10th edition was documented in the year prior to imaging: Soft tissue disorders (ICD10 Codes M60 - M79) |
| Chapter_XIII_5 | ICD10 | Osteopathies and chondropathies | Number of times an International Classification of Diseases, 10th edition was documented in the year prior to imaging: Osteopathies and chondropathies (ICD10 Codes M80 - M94) |
| Chapter_XIII_6 | ICD10 | Other disorders of the musculoskeletal system and connective tissue | Number of times an International Classification of Diseases, 10th edition was documented in the year prior to imaging: Other disorders of the musculoskeletal system and connective tissue (ICD10 Codes M95 - M99) |
| Chapter_XIV_1 | ICD10 | Glomerular diseases | Number of times an International Classification of Diseases, 10th edition was documented in the year prior to imaging: Glomerular diseases (ICD10 Codes N00 - N08) |
| Chapter_XIV_10 | ICD10 | Noninflammatory disorders of female genital tract | Number of times an International Classification of Diseases, 10th edition was documented in the year prior to imaging: Noninflammatory disorders of female genital tract (ICD10 Codes N80 - N98) |
| Chapter_XIV_11 | ICD10 | Other disorders of the genitourinary system | Number of times an International Classification of Diseases, 10th edition was documented in the year prior to imaging: Other disorders of the genitourinary system (ICD10 Codes N99 - N99) |

| | | | |
|---|---|---|---|
| Chapter_XIV_2 | ICD10 | Renal tubulo-interstitial diseases | Number of times an International Classification of Diseases, 10th edition was documented in the year prior to imaging: Renal tubulo-interstitial diseases (ICD10 Codes N10 - N16) |
| Chapter_XIV_3 | ICD10 | Renal failure | Number of times an International Classification of Diseases, 10th edition was documented in the year prior to imaging: Renal failure (ICD10 Codes N17 - N19) |
| Chapter_XIV_4 | ICD10 | Urolithiasis | Number of times an International Classification of Diseases, 10th edition was documented in the year prior to imaging: Urolithiasis (ICD10 Codes N20 - N23) |
| Chapter_XIV_5 | ICD10 | Other disorders of kidney and ureter | Number of times an International Classification of Diseases, 10th edition was documented in the year prior to imaging: Other disorders of kidney and ureter (ICD10 Codes N25 - N29) |
| Chapter_XIV_6 | ICD10 | Other diseases of urinary system | Number of times an International Classification of Diseases, 10th edition was documented in the year prior to imaging: Other diseases of urinary system (ICD10 Codes N30 - N39) |
| Chapter_XIV_7 | ICD10 | Diseases of male genital organs | Number of times an International Classification of Diseases, 10th edition was documented in the year prior to imaging: Diseases of male genital organs (ICD10 Codes N40 - N51) |
| Chapter_XIV_8 | ICD10 | Disorders of breast | Number of times an International Classification of Diseases, 10th edition was documented in the year prior to imaging: Disorders of breast (ICD10 Codes N60 - N64) |

| | | | |
|---|---|---|---|
| Chapter_XIV_9 | ICD10 | Inflammatory diseases of female pelvic organs | Number of times an International Classification of Diseases, 10th edition was documented in the year prior to imaging: Inflammatory diseases of female pelvic organs (ICD10 Codes N70 - N77) |
| Chapter_XIX_1 | ICD10 | Injuries to the head | Number of times an International Classification of Diseases, 10th edition was documented in the year prior to imaging: Injuries to the head (ICD10 Codes S00 - S09) |
| Chapter_XIX_10 | ICD10 | Injuries to the ankle and foot | Number of times an International Classification of Diseases, 10th edition was documented in the year prior to imaging: Injuries to the ankle and foot (ICD10 Codes S90 - S99) |
| Chapter_XIX_11 | ICD10 | Injuries involving multiple body regions | Number of times an International Classification of Diseases, 10th edition was documented in the year prior to imaging: Injuries involving multiple body regions (ICD10 Codes T00 - T07) |
| Chapter_XIX_12 | ICD10 | Injuries to unspecified part of trunk, limb or body region | Number of times an International Classification of Diseases, 10th edition was documented in the year prior to imaging: Injuries to unspecified part of trunk, limb or body region (ICD10 Codes T08 - T14) |
| Chapter_XIX_13 | ICD10 | Effects of foreign body entering through natural orifice | Number of times an International Classification of Diseases, 10th edition was documented in the year prior to imaging: Effects of foreign body entering through natural orifice (ICD10 Codes T15 - T19) |

| | | | |
|---|---|---|---|
| Chapter_XIX_14 | ICD10 | Burns and corrosions | Number of times an International Classification of Diseases, 10th edition was documented in the year prior to imaging: Burns and corrosions (ICD10 Codes T20 - T32) |
| Chapter_XIX_16 | ICD10 | Poisoning by drugs, medicaments and biological substances | Number of times an International Classification of Diseases, 10th edition was documented in the year prior to imaging: Poisoning by drugs, medicaments and biological substances (ICD10 Codes T36 - T50) |
| Chapter_XIX_17 | ICD10 | Toxic effects of substances chiefly nonmedicinal as to source | Number of times an International Classification of Diseases, 10th edition was documented in the year prior to imaging: Toxic effects of substances chiefly nonmedicinal as to source (ICD10 Codes T51 - T65) |
| Chapter_XIX_18 | ICD10 | Other and unspecified effects of external causes | Number of times an International Classification of Diseases, 10th edition was documented in the year prior to imaging: Other and unspecified effects of external causes (ICD10 Codes T66 - T78) |
| Chapter_XIX_19 | ICD10 | Certain early complications of trauma | Number of times an International Classification of Diseases, 10th edition was documented in the year prior to imaging: Certain early complications of trauma (ICD10 Codes T79 - T79) |
| Chapter_XIX_2 | ICD10 | Injuries to the neck | Number of times an International Classification of Diseases, 10th edition was documented in the year prior to imaging: Injuries to the neck (ICD10 Codes S10 - S19) |

| | | | |
|---|---|---|---|
| Chapter_XIX_20 | ICD10 | Complications of surgical and medical care, not elsewhere classified | Number of times an International Classification of Diseases, 10th edition was documented in the year prior to imaging: Complications of surgical and medical care, not elsewhere classified (ICD10 Codes T80 - T88) |
| Chapter_XIX_3 | ICD10 | Injuries to the thorax | Number of times an International Classification of Diseases, 10th edition was documented in the year prior to imaging: Injuries to the thorax (ICD10 Codes S20 - S29) |
| Chapter_XIX_4 | ICD10 | Injuries to the abdomen, lower back, lumbar spine and pelvis | Number of times an International Classification of Diseases, 10th edition was documented in the year prior to imaging: Injuries to the abdomen, lower back, lumbar spine and pelvis (ICD10 Codes S30 - S39) |
| Chapter_XIX_5 | ICD10 | Injuries to the shoulder and upper arm | Number of times an International Classification of Diseases, 10th edition was documented in the year prior to imaging: Injuries to the shoulder and upper arm (ICD10 Codes S40 - S49) |
| Chapter_XIX_6 | ICD10 | Injuries to the elbow and forearm | Number of times an International Classification of Diseases, 10th edition was documented in the year prior to imaging: Injuries to the elbow and forearm (ICD10 Codes S50 - S59) |
| Chapter_XIX_7 | ICD10 | Injuries to the wrist and hand | Number of times an International Classification of Diseases, 10th edition was documented in the year prior to imaging: Injuries to the wrist and hand (ICD10 Codes S60 - S69) |

| | | | |
|---|---|---|---|
| Chapter_XIX_8 | ICD10 | Injuries to the hip and thigh | Number of times an International Classification of Diseases, 10th edition was documented in the year prior to imaging: Injuries to the hip and thigh (ICD10 Codes S70 - S79) |
| Chapter_XIX_9 | ICD10 | Injuries to the knee and lower leg | Number of times an International Classification of Diseases, 10th edition was documented in the year prior to imaging: Injuries to the knee and lower leg (ICD10 Codes S80 - S89) |
| Chapter_XV_1 | ICD10 | Pregnancy with abortive outcome | Number of times an International Classification of Diseases, 10th edition was documented in the year prior to imaging: Pregnancy with abortive outcome (ICD10 Codes O00 - O08) |
| Chapter_XV_2 | ICD10 | Oedema, proteinuria and hypertensive disorders in pregnancy, childbirth and the puerperium | Number of times an International Classification of Diseases, 10th edition was documented in the year prior to imaging: Oedema, proteinuria and hypertensive disorders in pregnancy, childbirth and the puerperium (ICD10 Codes O10 - O16) |
| Chapter_XV_3 | ICD10 | Other maternal disorders predominantly related to pregnancy | Number of times an International Classification of Diseases, 10th edition was documented in the year prior to imaging: Other maternal disorders predominantly related to pregnancy (ICD10 Codes O20 - O29) |
| Chapter_XV_4 | ICD10 | Maternal care related to the fetus and amniotic cavity and possible delivery problems | Number of times an International Classification of Diseases, 10th edition was documented in the year prior to imaging: Maternal care related to the fetus and amniotic cavity and possible delivery problems (ICD10 Codes O30 - O48) |

| | | | |
|---|---|---|---|
| Chapter_XV_5 | ICD10 | Complications of labour and delivery | Number of times an International Classification of Diseases, 10th edition was documented in the year prior to imaging: Complications of labour and delivery (ICD10 Codes O60 - O75) |
| Chapter_XV_6 | ICD10 | Delivery | Number of times an International Classification of Diseases, 10th edition was documented in the year prior to imaging: Delivery (ICD10 Codes O80 - O84) |
| Chapter_XV_7 | ICD10 | Complications predominantly related to the puerperium | Number of times an International Classification of Diseases, 10th edition was documented in the year prior to imaging: Complications predominantly related to the puerperium (ICD10 Codes O85 - O92) |
| Chapter_XV_8 | ICD10 | Other obstetric conditions, not elsewhere classified | Number of times an International Classification of Diseases, 10th edition was documented in the year prior to imaging: Other obstetric conditions, not elsewhere classified (ICD10 Codes O94 - O99) |
| Chapter_XVI_1 | ICD10 | Fetus and newborn affected by maternal factors and by complications of pregnancy, labour and delivery | Number of times an International Classification of Diseases, 10th edition was documented in the year prior to imaging: Fetus and newborn affected by maternal factors and by complications of pregnancy, labour and delivery (ICD10 Codes P00 - P04) |
| Chapter_XVI_10 | ICD10 | Other disorders originating in the perinatal period | Number of times an International Classification of Diseases, 10th edition was documented in the year prior to imaging: Other disorders originating in the perinatal period (ICD10 Codes P90 - P96) |

| | | | |
|---|---|---|---|
| Chapter_XVI_2 | ICD10 | Disorders related to length of gestation and fetal growth | Number of times an International Classification of Diseases, 10th edition was documented in the year prior to imaging: Disorders related to length of gestation and fetal growth (ICD10 Codes P05 - P08) |
| Chapter_XVI_4 | ICD10 | Respiratory and cardiovascular disorders specific to the perinatal period | Number of times an International Classification of Diseases, 10th edition was documented in the year prior to imaging: Respiratory and cardiovascular disorders specific to the perinatal period (ICD10 Codes P20 - P29) |
| Chapter_XVI_6 | ICD10 | Haemorrhagic and haematological disorders of fetus and newborn | Number of times an International Classification of Diseases, 10th edition was documented in the year prior to imaging: Haemorrhagic and haematological disorders of fetus and newborn (ICD10 Codes P50 - P61) |
| Chapter_XVI_7 | ICD10 | Transitory endocrine and metabolic disorders specific to fetus and newborn | Number of times an International Classification of Diseases, 10th edition was documented in the year prior to imaging: Transitory endocrine and metabolic disorders specific to fetus and newborn (ICD10 Codes P70 - P74) |
| Chapter_XVI_8 | ICD10 | Digestive system disorders of fetus and newborn | Number of times an International Classification of Diseases, 10th edition was documented in the year prior to imaging: Digestive system disorders of fetus and newborn (ICD10 Codes P75 - P78) |

| | | | |
|---|---|---|---|
| Chapter_XVII_1 | ICD10 | Congenital malformations of the nervous system | Number of times an International Classification of Diseases, 10th edition was documented in the year prior to imaging: Congenital malformations of the nervous system (ICD10 Codes Q00 - Q07) |
| Chapter_XVII_10 | ICD10 | Other congenital malformations | Number of times an International Classification of Diseases, 10th edition was documented in the year prior to imaging: Other congenital malformations (ICD10 Codes Q80 - Q89) |
| Chapter_XVII_11 | ICD10 | Chromosomal abnormalities, not elsewhere classified | Number of times an International Classification of Diseases, 10th edition was documented in the year prior to imaging: Chromosomal abnormalities, not elsewhere classified (ICD10 Codes Q90 - Q99) |
| Chapter_XVII_2 | ICD10 | Congenital malformations of eye, ear, face and neck | Number of times an International Classification of Diseases, 10th edition was documented in the year prior to imaging: Congenital malformations of eye, ear, face and neck (ICD10 Codes Q10 - Q18) |
| Chapter_XVII_3 | ICD10 | Congenital malformations of the circulatory system | Number of times an International Classification of Diseases, 10th edition was documented in the year prior to imaging: Congenital malformations of the circulatory system (ICD10 Codes Q20 - Q28) |
| Chapter_XVII_4 | ICD10 | Congenital malformations of the respiratory system | Number of times an International Classification of Diseases, 10th edition was documented in the year prior to imaging: Congenital malformations of the respiratory system (ICD10 Codes Q30 - Q34) |

| | | | |
|---|---|---|---|
| Chapter_XVII_5 | ICD10 | Cleft lip and cleft palate | Number of times an International Classification of Diseases, 10th edition was documented in the year prior to imaging: Cleft lip and cleft palate (ICD10 Codes Q35 - Q37) |
| Chapter_XVII_6 | ICD10 | Other congenital malformations of the digestive system | Number of times an International Classification of Diseases, 10th edition was documented in the year prior to imaging: Other congenital malformations of the digestive system (ICD10 Codes Q38 - Q45) |
| Chapter_XVII_7 | ICD10 | Congenital malformations of genital organs | Number of times an International Classification of Diseases, 10th edition was documented in the year prior to imaging: Congenital malformations of genital organs (ICD10 Codes Q50 - Q56) |
| Chapter_XVII_8 | ICD10 | Congenital malformations of the urinary system | Number of times an International Classification of Diseases, 10th edition was documented in the year prior to imaging: Congenital malformations of the urinary system (ICD10 Codes Q60 - Q64) |
| Chapter_XVII_9 | ICD10 | Congenital malformations and deformations of the musculoskeletal system | Number of times an International Classification of Diseases, 10th edition was documented in the year prior to imaging: Congenital malformations and deformations of the musculoskeletal system (ICD10 Codes Q65 - Q79) |
| Chapter_XVIII_1 | ICD10 | Symptoms and signs involving the circulatory and respiratory systems | Number of times an International Classification of Diseases, 10th edition was documented in the year prior to imaging: Symptoms and signs involving the circulatory and respiratory systems (ICD10 Codes R00 - R09) |

| | | | |
|---|---|---|---|
| Chapter_XVIII_10 | ICD10 | Abnormal findings on examination of urine, without diagnosis | Number of times an International Classification of Diseases, 10th edition was documented in the year prior to imaging: Abnormal findings on examination of urine, without diagnosis (ICD10 Codes R80 - R82) |
| Chapter_XVIII_11 | ICD10 | Abnormal findings on examination of other body fluids, substances and tissues, without diagnosis | Number of times an International Classification of Diseases, 10th edition was documented in the year prior to imaging: Abnormal findings on examination of other body fluids, substances and tissues, without diagnosis (ICD10 Codes R83 - R89) |
| Chapter_XVIII_12 | ICD10 | Abnormal findings on diagnostic imaging and in function studies, without diagnosis | Number of times an International Classification of Diseases, 10th edition was documented in the year prior to imaging: Abnormal findings on diagnostic imaging and in function studies, without diagnosis (ICD10 Codes R90 - R94) |
| Chapter_XVIII_13 | ICD10 | Ill-defined and unknown causes of mortality | Number of times an International Classification of Diseases, 10th edition was documented in the year prior to imaging: Ill-defined and unknown causes of mortality (ICD10 Codes R95 - R99) |
| Chapter_XVIII_2 | ICD10 | Symptoms and signs involving the digestive system and abdomen | Number of times an International Classification of Diseases, 10th edition was documented in the year prior to imaging: Symptoms and signs involving the digestive system and abdomen (ICD10 Codes R10 - R19) |

| | | | |
|---|---|---|---|
| Chapter_XVIII_3 | ICD10 | Symptoms and signs involving the skin and subcutaneous tissue | Number of times an International Classification of Diseases, 10th edition was documented in the year prior to imaging: Symptoms and signs involving the skin and subcutaneous tissue (ICD10 Codes R20 - R23) |
| Chapter_XVIII_4 | ICD10 | Symptoms and signs involving the nervous and musculoskeletal systems | Number of times an International Classification of Diseases, 10th edition was documented in the year prior to imaging: Symptoms and signs involving the nervous and musculoskeletal systems (ICD10 Codes R25 - R29) |
| Chapter_XVIII_5 | ICD10 | Symptoms and signs involving the urinary system | Number of times an International Classification of Diseases, 10th edition was documented in the year prior to imaging: Symptoms and signs involving the urinary system (ICD10 Codes R30 - R39) |
| Chapter_XVIII_6 | ICD10 | Symptoms and signs involving cognition, perception, emotional state and behaviour | Number of times an International Classification of Diseases, 10th edition was documented in the year prior to imaging: Symptoms and signs involving cognition, perception, emotional state and behaviour (ICD10 Codes R40 - R46) |
| Chapter_XVIII_7 | ICD10 | Symptoms and signs involving speech and voice | Number of times an International Classification of Diseases, 10th edition was documented in the year prior to imaging: Symptoms and signs involving speech and voice (ICD10 Codes R47 - R49) |

| | | | |
|---|---|---|---|
| Chapter_XVIII_8 | ICD10 | General symptoms and signs | Number of times an International Classification of Diseases, 10th edition was documented in the year prior to imaging: General symptoms and signs (ICD10 Codes R50 - R69) |
| Chapter_XVIII_9 | ICD10 | Abnormal findings on examination of blood, without diagnosis | Number of times an International Classification of Diseases, 10th edition was documented in the year prior to imaging: Abnormal findings on examination of blood, without diagnosis (ICD10 Codes R70 - R79) |
| Chapter_XX_1 | ICD10 | Accidents | Number of times an International Classification of Diseases, 10th edition was documented in the year prior to imaging: Accidents (ICD10 Codes V01 - X59) |
| Chapter_XX_2 | ICD10 | Intentional self-harm | Number of times an International Classification of Diseases, 10th edition was documented in the year prior to imaging: Intentional self-harm (ICD10 Codes X60 - X84) |
| Chapter_XX_3 | ICD10 | Assault | Number of times an International Classification of Diseases, 10th edition was documented in the year prior to imaging: Assault (ICD10 Codes X85 - Y09) |
| Chapter_XX_4 | ICD10 | Event of undetermined intent | Number of times an International Classification of Diseases, 10th edition was documented in the year prior to imaging: Event of undetermined intent (ICD10 Codes Y10 - Y34) |
| Chapter_XX_5 | ICD10 | Legal intervention and operations of war | Number of times an International Classification of Diseases, 10th edition was documented in the year prior to imaging: Legal intervention and operations of war (ICD10 Codes Y35 - Y36) |

| | | | |
|---|---|---|---|
| Chapter_XX_6 | ICD10 | Complications of medical and surgical care | Number of times an International Classification of Diseases, 10th edition was documented in the year prior to imaging: Complications of medical and surgical care (ICD10 Codes Y40 - Y84) |
| Chapter_XX_8 | ICD10 | Supplementary factors related to causes of morbidity and mortality classified elsewhere | Number of times an International Classification of Diseases, 10th edition was documented in the year prior to imaging: Supplementary factors related to causes of morbidity and mortality classified elsewhere (ICD10 Codes Y90 - Y98) |
| Chapter_XXI_1 | ICD10 | Persons encountering health services for examination and investigation | Number of times an International Classification of Diseases, 10th edition was documented in the year prior to imaging: Persons encountering health services for examination and investigation (ICD10 Codes Z00 - Z13) |
| Chapter_XXI_2 | ICD10 | Persons with potential health hazards related to communicable diseases | Number of times an International Classification of Diseases, 10th edition was documented in the year prior to imaging: Persons with potential health hazards related to communicable diseases (ICD10 Codes Z20 - Z29) |
| Chapter_XXI_3 | ICD10 | Persons encountering health services in circumstances related to reproduction | Number of times an International Classification of Diseases, 10th edition was documented in the year prior to imaging: Persons encountering health services in circumstances related to reproduction (ICD10 Codes Z30 - Z39) |

| | | | |
|---|---|---|---|
| Chapter_XXI_4 | ICD10 | Persons encountering health services for specific procedures and health care | Number of times an International Classification of Diseases, 10th edition was documented in the year prior to imaging: Persons encountering health services for specific procedures and health care (ICD10 Codes Z40 - Z54) |
| Chapter_XXI_5 | ICD10 | Persons with potential health hazards related to socioeconomic and psychosocial circumstances | Number of times an International Classification of Diseases, 10th edition was documented in the year prior to imaging: Persons with potential health hazards related to socioeconomic and psychosocial circumstances (ICD10 Codes Z55 - Z65) |
| Chapter_XXI_6 | ICD10 | Persons encountering health services in other circumstances | Number of times an International Classification of Diseases, 10th edition was documented in the year prior to imaging: Persons encountering health services in other circumstances (ICD10 Codes Z70 - Z76) |
| Chapter_XXI_7 | ICD10 | Persons with potential health hazards related to family and personal history and certain conditions influencing health status | Number of times an International Classification of Diseases, 10th edition was documented in the year prior to imaging: Persons with potential health hazards related to family and personal history and certain conditions influencing health status (ICD10 Codes Z80 - Z99) |
| exp_wt_lab_chol_hdl | Labs | HDL Cholesterol: exponentially weighted average | Exponentially weighted average of measurements or estimates of high density lipoprotein cholesterol during the year prior to abdominal scan [mg/dL] |

| | | | |
|---|---|---|---|
| exp_wt_lab_chol_ldl | Labs | LDL Cholesterol: exponentially weighted average | Exponentially weighted average of measurements or estimates of low density lipoprotein cholesterol during the year prior to abdominal scan [mg/dL] |
| exp_wt_lab_chol_total | Labs | Total Cholesterol: exponentially weighted average | Exponentially weighted average of measurements or estimates of total cholesterol during the year prior to abdominal scan [mg/dL] |
| exp_wt_lab_gluc | Labs | Serum glucose: exponentially weighted average | Exponentially weighted average of measurements of serum glucose during the year prior to abdominal scan [mg/dL] |
| exp_wt_lab_hba1c | Labs | HbA1c: exponentially weighted average | Exponentially weighted average of measurements of hemoglobin A1c during the year prior to abdominal scan [%] |
| exp_wt_lab_trig | Labs | Triglycerides: exponentially weighted average | Exponentially weighted average of measurements or estimates of triglycerides during the year prior to abdominal scan [mg/dL] |
| latest_value_chol_hdl | Labs | HDL Cholesterol: latest value | Latest available value of high density lipoprotein cholesterol during the year prior to abdominal scan [mg/dL] |
| latest_value_chol_ldl | Labs | LDL Cholesterol: latest value | Latest available value of low density lipoprotein cholesterol during the year prior to abdominal scan [mg/dL] |
| latest_value_chol_total | Labs | Total Cholesterol: latest value | Latest available value of total cholesterol during the year prior to abdominal scan [mg/dL] |
| latest_value_gluc | Labs | Serum glucose: latest value | Latest available value of serum glucose during the year prior to abdominal scan [mg/dL] |
| latest_value_hba1c | Labs | HbA1c: latest value | Latest available value of hemoglobin A1c during the year prior to abdominal scan [%] |

| | | | |
|---|---|---|---|
| latest_value_trig | Labs | Triglycerides: latest value | Latest available value of triglycerides during the year prior to abdominal scan [mg/dL] |
| num_times_taken_chol_hdl | Labs | HDL Cholesterol: times measured | Number of times high density lipoprotein cholesterol was measured during the year prior to abdominal scan |
| num_times_taken_chol_ldl | Labs | LDL Cholesterol: times measured | Number of times low density lipoprotein cholesterol was measured during the year prior to abdominal scan |
| num_times_taken_chol_total | Labs | Total Cholesterol: times measured | Number of times total cholesterol was measured during the year prior to abdominal scan |
| num_times_taken_gluc | Labs | Serum glucose: times measured | Number of times serum glucose was measured during the year prior to abdominal scan |
| num_times_taken_hba1c | Labs | HbA1c: times measured | Number of times hemoglobin A1c was measured during the year prior to abdominal scan |
| num_times_taken_trig | Labs | Triglycerides: times measured | Number of times triglycerides were measured during the year prior to abdominal scan |
| bmi | Other | Body Mass Index | Latest available measurement or estimation of body mass index in the year prior to abdominal scan [kg/m^2] |
| smoker | Other | Smoker | Whether it was documented that the patient is a smoker in the year prior to abdominal scan |
| exp_wt_lab_diastolic | Vitals | Diastolic blood pressure: exponentially weighted average | Exponentially weighted average of measurements diastolic blood pressure during the year prior to abdominal scan [mm Hg] |
| exp_wt_lab_hr | Vitals | Heart rate: exponentially weighted average | Exponentially weighted average of measurements heart rate during the year prior to abdominal scan [beats per minute] |

| | | | |
|---|---|---|---|
| exp_wt_lab_resp | Vitals | Respiratory rate: exponentially weighted average | Exponentially weighted average of measurements respiratory rate during the year prior to abdominal scan [cycles per minute] |
| exp_wt_lab_spo2 | Vitals | SpO2: exponentially weighted average | Exponentially weighted average of measurements of oxygen saturation (SpO2) during the year prior to abdominal scan [%] |
| exp_wt_lab_systolic | Vitals | Systolic blood pressure: exponentially weighted average | Exponentially weighted average of measurements of systolic blood pressure during the year prior to abdominal scan [mm Hg] |
| exp_wt_lab_temp_c | Vitals | Temperature: exponentially weighted average | Exponentially weighted average of measurements temperature during the year prior to abdominal scan [Celsius] |
| latest_value_diastolic | Vitals | Diastolic blood pressure: latest value | Latest available value of diastolic blood pressure during the year prior to abdominal scan [mm Hg] |
| latest_value_hr | Vitals | Heart rate: latest value | Latest available value of heart rate during the year prior to abdominal scan [beats per minute] |
| latest_value_resp | Vitals | Respiratory rate: latest value | Latest available value of respiratory rate during the year prior to abdominal scan [cycles per minute] |
| latest_value_spo2 | Vitals | SpO2: latest value | Latest available value of oxygen saturation (SpO2) during the year prior to abdominal scan [%] |
| latest_value_systolic | Vitals | Systolic blood pressure: latest value | Latest available value of systolic blood pressure during the year prior to abdominal scan [mm Hg] |
| latest_value_temp_c | Vitals | Temperature: latest value | Latest available value of temperature during the year prior to abdominal scan [Celsius] |
| num_times_measured_diastolic | Vitals | Diastolic blood pressure: times measured | Number of times diastolic blood pressure was measured during the year prior to abdominal scan |

| | | | |
|---|---|---|---|
| num_times_measured_hr | Vitals | Heart rate: times measured | Number of times heart rate was measured during the year prior to abdominal scan |
| num_times_measured_resp | Vitals | Respiratory rate: times measured | Number of times respiratory rate was measured during the year prior to abdominal scan |
| num_times_measured_spo2 | Vitals | SpO2: times measured | Number of times oxygen saturation (SpO2) was measured during the year prior to abdominal scan |
| num_times_measured_systolic | Vitals | Systolic blood pressure: times measured | Number of times systolic blood pressure was measured during the year prior to abdominal scan |
| num_times_measured_temp_c | Vitals | Temperature: times measured | Number of times temperature was measured during the year prior to abdominal scan |
