## Supplemental Table 5 for "Opportunistic Assessment of Ischemic Heart Disease Risk Using Abdominopelvic Computed Tomography and Medical Record Data: a Multimodal Explainable Artificial Intelligence Approach"

| Subpopulation | Model | 1-year cohort | | 5-year cohort | |
|---|---|---|---|---|---|
| | | AUROC (95% CI) | AUCPR (95% CI) | AUROC (95% CI) | AUCPR (95% CI) |
| Complete PCE data (1y n=313, % IHD positive=4.5) / (5y n=72, % IHD positive=26.4) | FRS | .71 (.58-.82) | .09 (.07-.17) | .54 (.40-.65) | .28 (.24-.39) |
| | PCE | **.75 (.61-.85)** | **.17 (.09-.37)** | .60 (.46-.70) | .31 (.27-.43) |
| | Segmentation only | .67 (.55-.76) | .07 (.06-.11) | .56 (.42-.69) | .33 (.26-.50) |
| | PCE+Segmentation | .73 (.58-.85) | .12 (.08-.20) | .56 (.42-.68) | .31 (.26-.44) |
| | Clinical only | .63 (.51-.76) | .07 (.05-.14) | .78 (.66-.87) | .61 (.46-.79) |
| | Imaging only | .69 (.59-.82) | .09 (.06-.18) | .71 (.58-.80) | .51 (.37-.66) |
| | Imaging+Clinical Fusion | .67 (.57-.79) | .08 (.06-.17) | .79 (.67-.87) | .62 (.49-.78) |
| | Imaging+Clinical+Segmentation Fusion | .64 (.55-.77) | .08 (.05-.17) | **.81 (.69-.89)** | **.64 (.50-.81)** |
| Missing PCE data (1y n=1305, % IHD positive=4.4) / (5y n=282, % IHD positive=24.8) | FRS | .72 (.67-.77) | .09 (.07-.13) | .76 (.70-.81) | .49 (.41-.58) |
| | PCE | .75 (.71-.81) | .12 (.10-.18) | .77 (.72-.82) | .48 (.41-.57) |
| | Segmentation | .70 (.65-.75) | .08 (.07-.11) | .77 (.72-.82) | .46 (.40-.56) |
| | PCE+Segmentation | .77 (.73-.82) | .12 (.09-.17) | .79 (.74-.83) | .48 (.41-.58) |
| | Clinical only | **.80 (.76-.84)** | .16 (.12-.24) | .86 (.81-.89) | .67 (.60-.75) |
| | Imaging only | .75 (.71-.80) | .11 (.09-.16) | .83 (.78-.87) | .69 (.61-.77) |
| | Imaging+Clinical Fusion | .79 (.75-.84) | **.17 (.12-.24)** | **.88 (.84-.91)** | **.73 (.66-.81)** |
| | Imaging+Clinical+Segmentation Fusion | .77 (.72-.82) | **.17 (.12-.24)** | .87 (.83-.91) | .72 (.65-.80) |
| 40-75 y/o (1y n=1047, % IHD positive=4.5)/ (5y n=241, % IHD positive=27.8) | FRS | .63 (.57-.69) | .07 (.05-.09) | .66 (.59-.72) | .39 (.34-.47) |
| | PCE | .69 (.62-.75) | .08 (.07-.11) | .70 (.64-.76) | .43 (.37-.52) |
| | Segmentation | .67 (.61-.72) | .07 (.06-.10) | .68 (.62-.74) | .40 (.35-.49) |
| | PCE+Segmentation | .71 (.64-.76) | .10 (.08-.15) | .70 (.64-.76) | .41 (.36-.50) |
| | Clinical only | .70 (.64-.77) | .11 (.08-.17) | .84 (.79-.88) | .68 (.60-.77) |
| | Imaging only | .70 (.65-.77) | .10 (.08-.17) | .77 (.71-.82) | .63 (.56-.72) |
| | Imaging+Clinical Fusion | **.73 (.68-.80)** | **.13 (.09-.20)** | **.86 (.82-.90)** | **.73 (.66-.81)** |
| | Imaging+Clinical+Segmentation Fusion | .70 (.64-.78) | .12 (.09-.20) | .85 (.81-.89) | **.73 (.65-.81)** |
| Under 40 y/o (1y n=450, % IHD positive=1.1) / (5y n=81, % IHD positive=8.6) | FRS | .35 (.10-.38) | .01 (.01-.02) | .64 (.32-.83) | .29 (.12-.61) |
| | PCE | .46 (.12-.64) | .01 (.01-.02) | .76 (.59-.86) | .20 (.14-.43) |
| | Segmentation | .65 (.33-.84) | **.05 (.01-.19)** | .59 (.19-.84) | .32 (.12-.61) |
| | PCE+Segmentation | .56 (.23-.69) | **.05 (.01-.22)** | .70 (.39-.90) | .28 (.14-.60) |
| | Clinical only | **.69 (.29-.94)** | **.05 (.02-.15)** | .76 (.44-.95) | .40 (.20-.69) |
| | Imaging only | **.69 (.41-.94)** | .03 (.02-.08) | **.82 (.54-.97)** | .47 (.28-.84) |

| | | | | | |
|---|---|---|---|---|---|
| | Imaging+Clinical Fusion | **.69 (.33-.94)** | .04 (.02-.10) | .80 (.52-.97) | .51 (.27-.81) |
| | Imaging+Clinical+Segmentation Fusion | .68 (.43-.90) | .03 (.02-.07) | .80 (.49-.98) | **.55 (.31-.86)** |
| Over 75 y/o (1y n=121, % IHD positive=15.7) / (5y n=32, % IHD positive=46.9) | FRS | .47 (.33-.56) | .15 (.13-.22) | .51 (.36-.70) | .55 (.43-.73) |
| | PCE | .51 (.35-.61) | .19 (.14-.34) | .36 (.22-.55) | .40 (.36-.53) |
| | Segmentation | .43 (.30-.54) | .15 (.13-.24) | .64 (.45-.79) | .65 (.52-.82) |
| | PCE+Segmentation | .54 (.39-.65) | .19 (.15-.27) | .49 (.30-.65) | .49 (.40-.68) |
| | Clinical only | **.58 (.44-.68)** | **.22 (.17-.36)** | .61 (.42-.77) | .66 (.52-.80) |
| | Imaging only | .50 (.38-.63) | .17 (.14-.28) | **.76 (.61-.89)** | **.78 (.66-.89)** |
| | Imaging+Clinical Fusion | .54 (.39-.65) | .20 (.15-.33) | .70 (.52-.84) | .75 (.62-.88) |
| | Imaging+Clinical+Segmentation Fusion | .56 (.40-.67) | **.22 (.16-.37)** | .67 (.49-.82) | .72 (.59-.85) |
| Male (1y n=684, % IHD positive=5.4) / (5y n=125, % IHD positive=30.4) | FRS | .70 (.65-.77) | .11 (.08-.17) | .73 (.66-.81) | .48 (.41-.60) |
| | PCE | .74 (.69-.82) | **.15 (.11-.24)** | .73 (.64-.80) | .47 (.40-.60) |
| | Segmentation | .69 (.62-.74) | .09 (.08-.13) | .71 (.62-.78) | .46 (.39-.60) |
| | PCE+Segmentation | .73 (.67-.80) | .12 (.09-.18) | .74 (.65-.81) | .49 (.41-.61) |
| | Clinical only | **.76 (.71-.81)** | .13 (.10-.21) | .82 (.75-.88) | .69 (.60-.80) |
| | Imaging only | .72 (.66-.78) | .11 (.09-.18) | .84 (.77-.90) | .74 (.64-.83) |
| | Imaging+Clinical Fusion | .75 (.70-.81) | .14 (.10-.23) | **.88 (.82-.93)** | **.76 (.67-.85)** |
| | Imaging+Clinical+Segmentation Fusion | .72 (.67-.80) | **.15 (.11-.24)** | **.88 (.82-.92)** | .75 (.66-.85) |
| Female (1y n=934, % IHD positive=3.6) / (5y n=229, % IHD positive=22.3) | FRS | .76 (.69-.82) | .09 (.07-.15) | .73 (.66-.79) | .39 (.33-.50) |
| | PCE | .75 (.67-.82) | .11 (.08-.19) | .73 (.67-.79) | .38 (.32-.48) |
| | Segmentation | .73 (.66-.79) | .08 (.06-.12) | .73 (.66-.79) | .42 (.34-.53) |
| | PCE+Segmentation | **.78 (.70-.85)** | .12 (.09-.18) | .74 (.68-.80) | .40 (.33-.50) |
| | Clinical only | .75 (.67-.81) | .12 (.08-.20) | .84 (.78-.89) | .62 (.53-.72) |
| | Imaging only | .75 (.68-.81) | .10 (.07-.19) | .78 (.71-.84) | .57 (.48-.67) |
| | Imaging+Clinical Fusion | .77 (.69-.83) | **.13 (.09-.22)** | **.85 (.79-.90)** | .65 (.56-.75) |
| | Imaging+Clinical+Segmentation Fusion | .74 (.66-.81) | .12 (.09-.21) | .84 (.79-.89) | **.66 (.57-.75)** |
| Asian (1y n=257, % IHD positive=4.3) / (5y n=55, % IHD positive=32.7) | FRS | .73 (.64-.81) | .08 (.07-.14) | .73 (.62-.86) | .59 (.46-.75) |
| | PCE | **.82 (.74-.89)** | .15 (.10-.34) | **.78 (.66-.89)** | **.66 (.52-.81)** |
| | Segmentation | .65 (.56-.74) | .06 (.05-.10) | .77 (.65-.89) | .64 (.51-.81) |
| | PCE+Segmentation | .80 (.72-.87) | .11 (.08-.19) | .76 (.64-.88) | .61 (.48-.78) |
| | Clinical only | .81 (.73-.88) | .14 (.10-.29) | .72 (.60-.84) | .56 (.43-.74) |
| | Imaging only | .76 (.66-.85) | .11 (.08-.21) | .76 (.64-.90) | .72 (.60-.85) |

| | | | | | |
|---|---|---|---|---|---|
| | Imaging+Clinical Fusion | .81 (.74-.88) | .15 (.10-.35) | .75 (.63-.88) | **.66 (.53-.81)** |
| | Imaging+Clinical+Segmentation Fusion | .81 (.74-.88) | **.18 (.10-.38)** | .74 (.63-.86) | .62 (.48-.77) |
| Non-Hispanic Black (1y n=64, % IHD positive=3.1)/ (5y n=13, % IHD positive=23.1) | FRS | .92 (.88-.98) | .23 (.15-.50) | .77 (.25-1.00) | .77 (.50-1.00) |
| | PCE | .83 (.75-.93) | .12 (.09-.25) | .73 (.36-1.00) | .66 (.32-1.00) |
| | Segmentation | **.97 (.92-1.00)** | **.42 (.27-1.00)** | .73 (.36-1.00) | .66 (.31-1.00) |
| | PCE+Segmentation | .94 (.88-.98) | .33 (.22-.67) | .97 (.83-1.00) | .92 (.73-1.00) |
| | Clinical only | .76 (.53-.97) | .14 (.06-.50) | .93 (.75-1.00) | .81 (.53-1.00) |
| | Imaging only | .94 (.89-.98) | .29 (.19-.67) | .70 (.17-1.00) | .65 (.29-1.00) |
| | Imaging+Clinical Fusion | .90 (.85-.97) | .18 (.13-.40) | .93 (.75-1.00) | .87 (.67-1.00) |
| | Imaging+Clinical+Segmentation Fusion | .83 (.74-.94) | .12 (.09-.29) | **1.00 (1.00-1.00)** | **1.00 (1.00-1.00)** |
| Hispanic (1y n=306, % IHD positive=4.9)/ (5y n=79, % IHD positive=27.8) | FRS | .74 (.61-.86) | **.18 (.10-.37)** | .83 (.74-.92) | .68 (.55-.83) |
| | PCE | .71 (.58-.82) | **.18 (.09-.38)** | .87 (.79-.94) | .67 (.55-.83) |
| | Segmentation | .68 (.57-.79) | .11 (.07-.23) | .78 (.69-.86) | .52 (.41-.67) |
| | PCE+Segmentation | .77 (.68-.85) | .14 (.10-.25) | .83 (.75-.91) | .61 (.49-.77) |
| | Clinical only | **.77 (.70-.84)** | .11 (.09-.19) | **.90 (.80-.97)** | **.83 (.72-.93)** |
| | Imaging only | .71 (.60-.80) | .11 (.07-.21) | .78 (.69-.87) | .56 (.44-.71) |
| | Imaging+Clinical Fusion | .73 (.64-.82) | .13 (.08-.29) | .89 (.81-.95) | .80 (.69-.90) |
| | Imaging+Clinical+Segmentation Fusion | .70 (.60-.80) | .12 (.08-.28) | .87 (.78-.95) | .78 (.66-.89) |
| Other (1y n=189, % IHD positive=3.7) / (5y n=32, % IHD positive=25.0) | FRS | .65 (.58-.73) | .06 (.05-.08) | .43 (.27-.61) | .23 (.21-.35) |
| | PCE | **.81 (.66-.92)** | .12 (.08-.26) | .46 (.29-.63) | .24 (.21-.37) |
| | Segmentation | .60 (.46-.74) | .05 (.04-.08) | .69 (.53-.86) | .42 (.31-.70) |
| | PCE+Segmentation | .72 (.61-.82) | .08 (.06-.13) | .48 (.32-.64) | .24 (.22-.36) |
| | Clinical only | .77 (.63-.89) | .12 (.07-.30) | .64 (.41-.81) | .53 (.33-.76) |
| | Imaging only | .78 (.68-.88) | .09 (.07-.18) | **.90 (.74-1.00)** | **.86 (.70-1.00)** |
| | Imaging+Clinical Fusion | **.81 (.69-.91)** | .12 (.08-.25) | .77 (.58-.90) | .63 (.43-.84) |
| | Imaging+Clinical+Segmentation Fusion | .80 (.62-.93) | **.14 (.09-.29)** | .75 (.54-.91) | .65 (.45-.86) |
| Non-Hispanic White (1y n=802, % IHD positive=4.5) / (5y n=175, % IHD positive=21.7) | FRS | .69 (.64-.77) | .10 (.07-.18) | .70 (.62-.76) | .32 (.27-.42) |
| | PCE | .75 (.70-.83) | .13 (.09-.22) | .70 (.62-.77) | .34 (.29-.45) |
| | Segmentation | .72 (.66-.78) | .09 (.07-.13) | .68 (.59-.75) | .35 (.29-.47) |
| | PCE+Segmentation | .75 (.69-.83) | .15 (.11-.24) | .72 (.64-.79) | .36 (.31-.49) |
| | Clinical only | .75 (.69-.83) | .18 (.11-.29) | .85 (.79-.90) | .62 (.51-.73) |

| | | | | | |
|---|---|---|---|---|---|
| | Imaging only | .73 (.66-.80) | .14 (.09-.23) | .85 (.79-.90) | .65 (.55-.76) |
| | Imaging+Clinical Fusion | **.77 (.70-.84)** | **.19 (.13-.31)** | **.90 (.85-.93)** | .69 (.59-.80) |
| | Imaging+Clinical+Segmentation Fusion | .73 (.65-.81) | **.19 (.12-.31)** | **.90 (.85-.94)** | **.72 (.63-.83)** |
| Taking lipid modifying agents (1y n=375, % IHD positive=7.7) / (5y n=105, % IHD positive=37.1) | FRS | .72 (.65-.79) | .14 (.12-.20) | .69 (.60-.77) | .50 (.43-.62) |
| | PCE | .72 (.66-.80) | .17 (.13-.26) | .70 (.62-.78) | .50 (.43-.61) |
| | Segmentation | .65 (.59-.72) | .11 (.09-.17) | .66 (.57-.75) | .53 (.44-.65) |
| | PCE+Segmentation | **.76 (.68-.82)** | .18 (.14-.24) | .71 (.63-.79) | .50 (.44-.63) |
| | Clinical only | .73 (.64-.80) | .19 (.14-.30) | .85 (.78-.91) | .79 (.71-.88) |
| | Imaging only | .71 (.63-.80) | .16 (.12-.25) | .83 (.75-.90) | .79 (.71-.86) |
| | Imaging+Clinical Fusion | **.76 (.68-.83)** | **.22 (.16-.35)** | .88 (.81-.93) | .83 (.76-.91) |
| | Imaging+Clinical+Segmentation Fusion | .74 (.64-.82) | **.22 (.16-.35)** | **.89 (.82-.94)** | **.86 (.79-.92)** |
| Not taking lipid modifying agents (1y n=1243, % IHD positive=3.4) / (5y n=249, % IHD positive=20.1) | FRS | .67 (.61-.74) | .06 (.05-.11) | .69 (.62-.75) | .31 (.27-.40) |
| | PCE | .73 (.68-.81) | **.10 (.07-.18)** | .72 (.65-.77) | .33 (.29-.43) |
| | Segmentation | .69 (.62-.75) | .06 (.05-.09) | .73 (.66-.79) | .37 (.31-.48) |
| | PCE+Segmentation | .73 (.67-.79) | .08 (.06-.11) | .73 (.66-.78) | .36 (.29-.45) |
| | Clinical only | .75 (.70-.81) | .08 (.07-.12) | .83 (.77-.87) | .51 (.42-.62) |
| | Imaging only | .74 (.70-.80) | .08 (.06-.14) | .78 (.71-.83) | .51 (.42-.62) |
| | Imaging+Clinical Fusion | **.76 (.71-.81)** | .09 (.07-.13) | **.85 (.79-.89)** | **.54 (.46-.66)** |
| | Imaging+Clinical+Segmentation Fusion | .72 (.67-.79) | .08 (.06-.13) | .84 (.79-.88) | .53 (.44-.65) |
